# Effectiveness of neurofeedback interventions in cerebral palsy: A systematic review

**DOI:** 10.1101/2025.08.14.25333687

**Authors:** Sevinc Nisa Abay, Kaat Alaerts, Bernard Dan, Elegast Monbaliu, Saranda Bekteshi

**Author notes:** Corresponding author: Dr. Saranda Bekteshi, KU Leuven, Department of Rehabilitation Sciences, Neurorehabilitation Technology Lab.

## Abstract

**Aim:** To synthesise existing evidence on the effectiveness of neurofeedback (NF) interventions in individuals with cerebral palsy (CP).

**Method:** A systematic search was conducted in nine databases in October 2023, December 2023, and November 2024, without language restrictions. Studies were eligible if they investigated NF interventions in individuals with CP, irrespective of intervention type, study design, or participant characteristics (e.g. diagnosis, sex, age). Study quality was appraised using the Downs and Black checklist and the Single-Case Experimental Design (SCED) Scale. Risk of bias was assessed with the revised Cochrane risk-of-bias tool (RoB 2), the Cochrane Risk of Bias in Non-randomised Studies of Intervention (ROBINS-I V2), and the Single-Case Design RoB tool.

**Results:** Eighteen studies (n=439, age range 4-47 years) were included: seven randomised controlled trials (RCTs), seven non-RCTs, and four SCED studies. All interventions used electroencephalography (EEG)-based NF, with some integrating brain–computer interfaces (BCIs) or external devices such as powered wheelchairs and robotic hands. Reported benefits included improvements in brain activity, motor outcomes, and cognitive functions. However, not all studies confirmed these effects, and findings were inconsistent across protocols. The use of multiple tools and external devices appeared to strengthen intervention effects in some cases.

**Interpretation:** NF interventions in CP show potential, but their effectiveness remains inconclusive, largely due to heterogeneity in intervention protocols (i.e., frequency bands targeted, electrode placement, feedback modalities, and intervention intensity) and limited follow-up data. Future studies should prioritise rigorously designed RCTs, incorporate personalised approaches, and consider multimodal NF interventions.

**What this paper adds:**

- Individuals with cerebral palsy are capable of learning to regulate their neural activity through neurofeedback.
- Neurofeedback interventions show potential to improve motor, cognitive, and functional outcomes, although findings remain inconsistent.
- The use of brain–computer interfaces and external devices may enhance the effectiveness of neurofeedback.
- Protocol heterogeneity and limited follow-up remain key barriers to establishing neurofeedback effectiveness.

## INTRODUCTION

Cerebral palsy (CP) is an early-onset lifelong neurodevelopmental condition with a prevalence of 1.6-3.4 per 1000 live births, characterised by limitations in activity due to impaired development of movement and posture.^1, 2^ CP is categorised into spastic, dyskinetic, ataxic and mixed forms, and the functioning of individuals ranges from mild to severe levels of limitations.^3^ Outcomes of CP can be categorised under the International Classification of Functioning, Disability and Health (ICF) body function and structure, and activity and participation domains.^4^ Within the ICF framework, impairments in body functions and structures associated with CP primarily affect motor functioning. Out of all children with CP, 40% are unable to walk independently,^5^ 23% experience difficulties in hand functioning,^6^ and 32% in speech intelligibility.^7^ This profile represents the more severe cases of CP, who rely on assistive technologies for optimal daily life functioning, including powered wheelchairs for mobility and eye-tracking technology for communication and computer access.^8^ In addition to this broad range of impairments, 95% of individuals with CP have at least one comorbidity, while 36.4% experience at least one disorder within neurological, medical and mental disorder categories.^9^ Consequently, within the ICF activity and participation domain, CP impacts individuals’ quality of life (QoL) and societal participation.^10^ Improving daily life functioning of individuals with CP requires an integrative approach including physical therapy, occupational therapy, speech-language therapy, medical and surgical treatments, and teleinterventions.^8^

Self-regulated neural plasticity is increasingly recognised as a key driver of lasting motor and cognitive gains in rehabilitation.^11^ Neurofeedback (NF) operationalises this principle as a training method that enables self-regulation of brain activity by providing real-time feedback of neurophysiological signals recorded from the brain.^12, 13^ Brain activity can be measured through functional magnetic resonance imaging (fMRI), electroencephalogram (EEG), magnetoencephalography (MEG), and functional near-infrared spectroscopy (fNIRS) in order to provide real-time information of neural activity.^14^ This non-invasive training method aims to practice self-regulation of the neural foundation that underlies a pathology or dysfunction.^13^ NF training is often conducted integrating brain-computer interface (BCI), a technology that converts brain activity into digital commands, enabling direct communication between the brain and external devices for applications ranging from assistive technologies to interactive entertainment.^15^ BCI technology has been used with children with CP, proving feasible in controlling wheelchairs^16^ and computer access.^17, 18^ BCI can be particularly useful for children with severe CP who lack motor control and coordination,^19^ providing them with the possibility to interact independently with their environment.

The effectiveness of NF training in healthy^20, 21^ and clinical populations^22^ has been widely researched and synthesised in various systematic reviews, including Attention Deficit Hyperactivity Disorder (ADHD),^23, 24^ Autism Spectrum Disorder (ASD),^25, 26^ stroke,^27^ brain injury,^28^ Parkinson’s,^29^ Post-Traumatic Stress Disorder,^30^ and pain management.^31^ NF is a complex treatment approach with its pros and cons, and to date, scientific evidence to support its implementation remains inconclusive.^32^

In the field of CP, there is a growing interest in NF as a non-invasive neurorehabilitation approach, with numerous studies published in recent years. Synthesising the existing evidence of its effectiveness in the goal group will inform clinical practice and guide future research.

To address a critical gap in the literature, this systematic review aims to synthesise current evidence on NF interventions in individuals with CP. It focuses on examining the methodological rigour and therapeutic impact of these interventions across domains of functioning, as defined by the ICF psychosocial framework. By critically evaluating existing studies, this review seeks to guide the development of more targeted and effective NF-based approaches, ultimately supporting clinicians in optimising therapeutic outcomes for people with CP.

## METHOD

This systematic review was organised and reported according to Preferred Reporting Items for Systematic Reviews and Meta-Analyses (PRISMA)^33, 34^ guidelines and was registered with PROSPERO (CRD42023473422). A change regarding the review timeline was made to the registered protocol on 27 November 2023; the end date was changed from 31 May 2024 to 31 January 2025. For the critical appraisal of this review, the A MeaSurement Tool to Assess systematic Reviews 2 (AMSTAR-2)^35^ checklist was used.

## Search strategy

In the context of this review, three literature searches were conducted by the first author (SNA) with supervision of the last author (SB). Nine electronic databases were searched, including Embase, PubMed, PsycINFO, MEDLINE (Ovid), Scopus, CINAHL (EBSCO), ProQuest, Web of Science, and Cochrane Library without any restriction on the publication language.

The first search was conducted from intercept to 26 October 2023 with the search strategy; ((“neurofeedback” OR “neurotherap*” OR “eeg biofeedback” OR “brain wave”) AND “cerebral palsy”).

With the agreement of all authors to be more inclusive and include papers about BCI, the second search was conducted on 24 December 2024 with the search strategy ((((“brain-computer interface”) OR (“brain computer interface”)) OR (“brain-machine interface”)) OR (“brain machine interface”)) AND (“cerebral palsy”).

The third search was conducted on 19 November 2024, covering the period between October 2023 to October 2024, using the following search strategy; ((“brain-computer interface” OR “brain computer interface” OR “brain-machine interface” OR “brain machine interface” OR neurofeedback OR “eeg biofeedback” OR neurotherap* OR “brain wave”) AND “cerebral palsy”) AND 2023/10/26:2024/10/19. The three search strategies used in each database are shown in Appendix S1.

Deduplication was done using EndNote X9 (Clarivate Analytics, Philadelphia, PA, USA). Deduplicated records were then imported into Rayyan-Qatar Computing Institute Research^36^ for screening and labelling.

### Inclusion and exclusion criteria

The inclusion criteria were: (1) targeting individuals diagnosed with CP, with no restriction on the type of CP, age of participant, and severity of symptoms; (2) intervention studies (non-randomised and randomised) focused on NF training in individuals with CP. The study should have at least a pre-post assessment to be eligible. The NF intervention studies may focus on any goals (such as upper-lower limb motor training, cognitive, activity and participation, and others); (3) any comparator (like non-exposure control groups) or lack of comparator; (4) peer-reviewed journal publications. To summarise, according to PICO principles, this systematic review focused on individuals with CP (P), receiving NF intervention (I), which may be compared to a non-exposure control group, or lack a comparator (C), aiming to improve outcomes of CP (O). The exclusion criteria were: (1) all studies focusing on populations other than CP, such as NF interventions in ASD or ADHD, (2) NF studies that are not experimental (i.e. do not include a pre-post study design), and (3) reviews and studies with a qualitative design.

### Selection procedure

Two authors (SNA) and (SB) completed the initial screening of titles and abstracts independently. All records were labelled as “exclude”, “include”, or “maybe” by both authors independently using separate password-protected Rayyan accounts. When there was a doubt about a study, it was included for full-text reading to prevent missing any relevant papers. Google Translate^37, 38^ was used to translate non-English abstracts and papers that were included for full-text reading. After initial screening, all results were discussed between both authors, and an agreement was reached regarding the records included for full-text review. The same process was repeated for full-text review records; after independently screening these records, both authors made decisions about the inclusion and exclusion of records. Any disagreement regarding these decisions was solved through a consensus meeting. Reference lists of all included papers were searched manually by both authors to extract any additional relevant papers.

### Data extraction

Data extraction was done using an Excel spreadsheets including study ID (title, author, and year), participant characteristics (diagnosis, diagnosis characteristics, clinical profile, sex, age), study characteristics (study design, having a control condition or not, details about control condition, sample size, baseline, follow up, outcome measurements), intervention details (device, sampling rate & impedance, aim of intervention, duration, interventionist, place of intervention, type of NF, interest of frequency) and results and conclusion. Data extraction was completed by one reviewer (SNA) and cross-checked by a second reviewer (SB). Full agreement was achieved through consensus meetings.

### Study quality appraisal and risk of bias assessment

Two authors (SNA and SB) independently conducted methodological quality appraisal and risk of bias assessments for each record included in this review; full agreement was reached through consensus.

Two different tools were used to assess the quality of evidence for the cohort studies and the single-case experimental design (SCED) studies, respectively. The modified version of the Downs and Black quality assessment tool^39^ was employed to assess the quality appraisal of the cohort studies. This assessment tool demonstrates high internal consistency, good test-retest and interrater reliability, and good face and criterion validity for non-randomised studies.^39^ This scale consists of 27 items across five subscales, including reporting (n=10), external validity (n=3), internal validity-bias (n=7), internal validity confounding (n=6), and power (n=1). While 25 of the items have a score of either 1 (representing acceptable quality) or 0 (representing not acceptable quality), item 5 can have a maximum score of 2 and item 27 can have scores ranging from 0 to 5. Therefore, the maximum score could be 32 or lower, while a higher score represents better methodological quality. Quality levels were used as reported by Hooper et al.,^40^ that is, excellent (26-32); good (20-25); fair (15-19); and poor (≤14). SCED scale^41^ was used to assess methodological appraisal of SCED studies. This scale has excellent interrater reliability^41, 42^ and consists of 11 items, including clinical history, target behaviours, study design, sufficient baseline measures, sampling behaviour during treatment, raw data recorded, interrater reliability, independence of assessors, statistical analysis, replication, and generalisation. While item 1 does not contribute to the general score, quality of items can be either 1 as yes (presence) and 0 as no (absence), scores ranged from 0 to 4 represents weak, scores ranged from 5 to 7 represents moderate and scores 8 to 10 represent high quality of evidence, score 10 being highest quality of evidence.

Risk of bias was assessed using three different tools, depending on the study design, that is, A revised Cochrane risk-of-bias tool (RoB 2)^43^ for Randomised Controlled Trials (RCTs), the Cochrane’s Risk of Bias in Non-randomised studies of Intervention (ROBINS-I V2)^44^ for non-RCTs, and the Single-Case Design RoB tool (SCD)^45^ for SCED studies.

RoB 2 consists of six domains: selection bias, performance bias, detection bias, attrition bias, reporting bias, and other bias. Each domain includes several items that can be rated as ‘yes,’ ‘partially yes,’ ‘partially no,’ or ‘no.’ Based on the responses to these items, the risk of bias judgment for each domain is categorised as low, high, or having some concerns, which then informs the overall risk of bias for the study.

ROBINS-I V2 (released November 2024) consists of seven domains: bias due to confounding, selection of participants into the study or into the analysis, classification of interventions, deviations from the intended interventions, missing data, measurement of outcomes and selection of the reported results. Each domain includes several items that can be answered as ‘yes’,‘partially yes’, ‘partially no’ or ‘no’; and for some questions ‘weak no’ and ‘strong no’. Based on the responses to these items, the risk of bias judgment for each domain is categorised as low, moderate, serious, or critical, which then determines the overall risk of bias for the study.

The SCD assessment tool was used to assess the risk of bias of SCED studies. This assessment tool included criteria such as selection bias, performance bias, and detection bias. The risk of bias for each criterion is labelled as low risk, unclear risk, or high risk.

### Data synthesis and reporting

Despite a considerable number of included publications, meta-analysis was deemed not feasible due to heterogeneity in NF protocols, study design, sample size, and outcome measures.^31, 46^ Therefore, this systematic review was conducted according to Synthesis Without Meta-analyses reporting guidelines as a formal narrative synthesis.^47^ Data were grouped based on study characteristics, participant characteristics, intervention characteristics, outcome measurement, and results. Outcomes of the NF intervention were reported across three domains: brain outcomes, motor outcomes, and cognitive outcomes. The effects of NF delivered via external devices and BCI-based NF were synthesised separately.

## RESULTS

### Search results

The details and steps of the database search are summarised in the PRISMA flow diagram in Figure 1. A total of 8,322 records were identified through the database search. After deduplication, 2,518 records were removed, leaving 5,804 records for title and abstract screening. Following this screening, 96 records were selected for full-text review. Of these, 78 records were excluded due to the absence of NF intervention, lack of participants with CP, or because the study was not peer-reviewed. The list of excluded records, along with the reasons for exclusion, is provided in Appendix S2. In total, 18 records were included in this systematic review. Fifteen studies were in English, two were in Russian, and one was in Portuguese.

**Figure 1.**
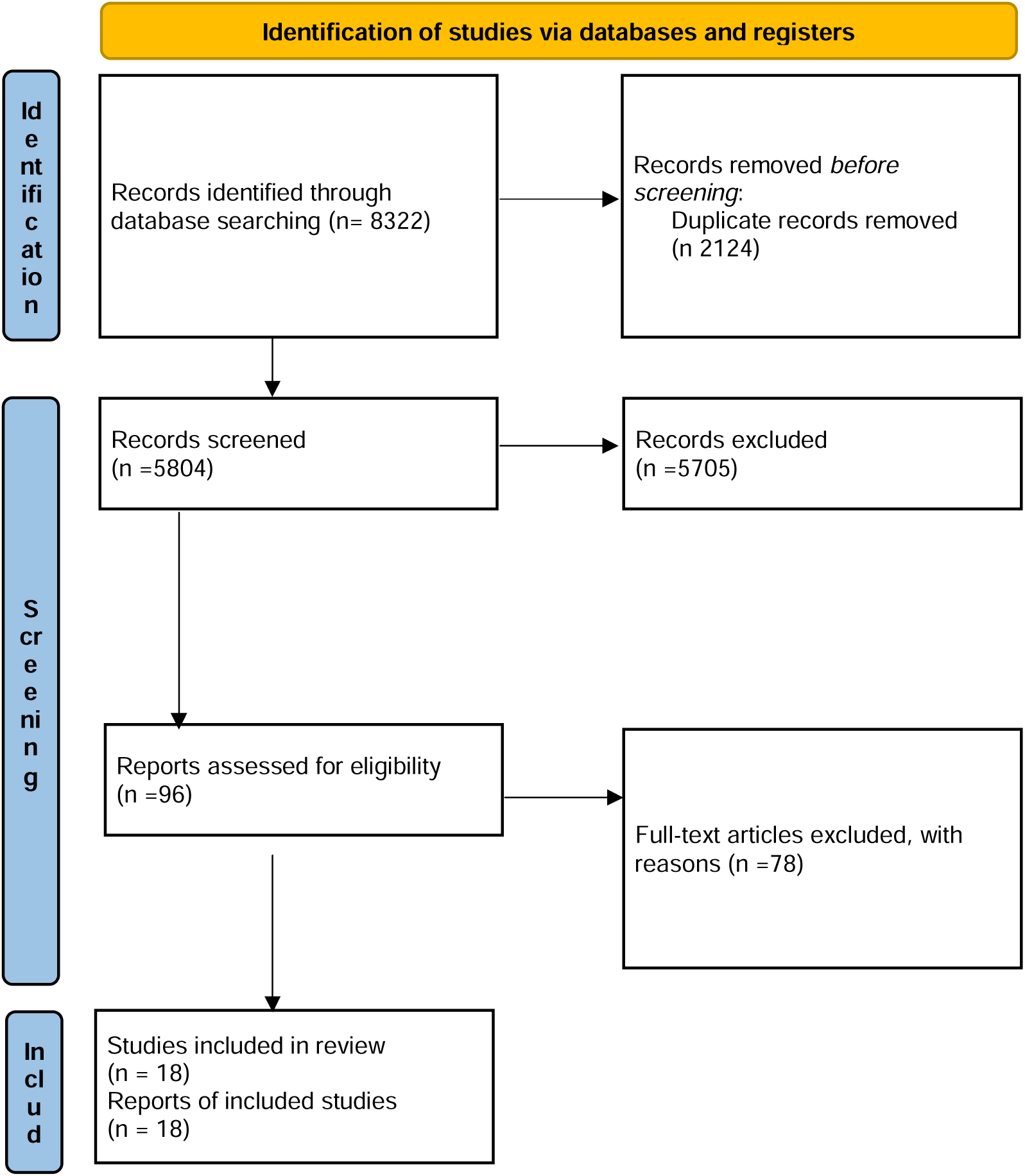
PRISMA diagram

### Participant characteristics

The 18 included studies comprised a total of 439 participants, including both individuals diagnosed with CP and healthy controls. The severity of CP ranged from mild to severe, as classified by the Gross Motor Function Classification System (GMFCS).^48^ Most studies included participants with mild CP; only four studies included individuals with severe CP. The age of participants ranged from 4 to 47 years. While the majority of studies focused on participants aged 6 to 18 years, five studies included adults over the age of 18. Participant characteristics are reported in Table 1.

**Table 1.** Summary of studies - Participant characteristics.

**Figure 1. PRISMA diagram**
| **Table 1. Summary of studies - Participant characteristics** |  |  |  |  |  |
| --- | --- | --- | --- | --- | --- |
| **Study** | **Diagnosis** | **Sample size, n** | **Sex (M:F)** | **Age** | **Clinical profile of CP** |
| Alves-Pinto et al., 2017 | Cerebral palsy | 11 | 5:6 | 21-47 | MACS – Level I to III<br>GMFCS – Level II to IV |
| Filho et al., 2017 | Cerebral palsy | 1 | 0:1 | 4 | MAS – 2 |
| Gupta et al., 2020 | Cerebral palsy | 60 | 34:26 | 5-15 | GMFCS – II and III |
| Kim & Lee., 2016 | Cerebral palsy | 18 | 11:7 | 4-9 | GMFCS – III and IV |
| Larina et al., 2020 | Cerebral palsy | 32 | 19:13 | 10-18 | GMFCS – >III |
| Legarda et al., 2011 | Cerebral palsy and autism<br>n=1, epilepsy n=2 | 3 | 2:1 | 6 -19 | Walking independently<br>with<br>bilateral ankle-foot<br>orthoses |
| Neuper et al., 2003 | Cerebral palsy | 1 | 1:0 | 32 | Severe spastic form of<br>tetraparesis and inability to<br>speak |
| Taherien et al., 2015 | Cerebral palsy | 8 | 4:4 | 7-43 | Spastic quadriplegic CP |
| Korandeh et al., 2021 | Cerebral palsy | 26 | NI | 4-10 | SCFMG – Level I and II |
| Yu et al., 2012 | Cerebral palsy | 28 | 17:11 | mean=9 | NI |
| Larina et al., 2019 | Cerebral palsy | 50 | 24:26 | 12-18 | GMFCS – ≤ III |
| Taherian et al., 2016 | Cerebral palsy | 1 | 0:1 | 21 | GMFCS – IV and V |
| Xie et al., 2021 | Cerebral palsy | 18 | 10:10 | 8-12 | NI |
| Korsunskaya et al., 2020 | Cerebral palsy | 30 | 36%:74% | 6-17 | NI |
| Pavlenko et al., 2023 | Cerebral palsy | 60 | 26:34 | 6-15 | GMFCS – not exceeding<br>III |
| Pavlenko et al., 2024 | Cerebral palsy | 50 | 35:15 | 7-15 | Spastic diplegia |
| Chen et al., 2024 | Cerebral palsy combined with AD | 19 | 17:2 | 3-6 | GMFCS and MACS – I - II |
| Pavlenko et al., 2024 | Cerebral palsy | 23 | 16:7 | 7-15 | GMFCS – I - III |
| CP: Cerebral Palsy, GMFCS: Gross Motor Function Classification System, MACS: Manual Ability Classification System, MAS: Modified Ashworth Scale, SCFMG: Shahid Chamran Functional Motor Grading, AD: Autism Disorder, NI: Not Indicated |  |  |  |  |  |

### Study characteristics

Of the 18 included studies, seven were RCTs,^49–55^ seven were non-RCTs,^56–62^ and four SCEDs.^17, 18, 63, 64^ Only one study included a follow-up assessment.^18^ Intervention protocols varied considerably across studies. The number of sessions ranged from a minimum of 3 to a maximum of 30. Sessions were typically conducted either daily with minimal breaks or twice a week with longer intervals between sessions. Session durations ranged from 8 to 45 minutes, with most studies implementing sessions of approximately 30 minutes.

In all studies, brain activity was measured using EEG, although the number of electrodes and sampling rates differed across studies. The type of feedback provided also varied: while most studies used visual feedback, some employed external devices to deliver feedback through BCI. The NF protocols targeted different EEG frequency bands, including reinforcement and/or inhibition of alpha rhythm, sensorimotor rhythm (SMR), high-beta, mid-beta, and infraslow oscillations. Instructions given to participants during NF training included mental tasks such as achieving a specific mental state to play a video, visualizing an action, or engaging in motor imagery. Study characteristics and intervention characteristics are presented in Table 2 and Table 3, respectively.

**Table 2.** Summary of studies - Study characteristics.

| **Table 2. Summary of studies - Study characteristics** |  |  |  |  |  |  |
| --- | --- | --- | --- | --- | --- | --- |
| **Study** | **Design** | **Baseline** | **Follow up** | **Control Condition** | **Outcome measures** | **Reported funding** |
| Alves-Pinto et al., 2017 | Non-randomized trial | EEG recording | NI | All participants attended - Simulated training | EEG and performance in SRTT | Buhl-Strohmaier Foundation and the Kraußianum Foundation |
| Filho et al., 2017 | Case report | EEG recording and MAS | NI | NA | EEG and MAS | NI |
| Gupta et al., 2020 | Randomized controlled trial | EEG recording and VSMS | NI | n=15 - only received standard therapy<br>n=15 - transcranial magnetic stimulation<br>n= 15 - cognitive attention exercises | VSMS and EEG | Department of Science and Technology (DST), Government of India, New Delhi to North-Eastern Hill University Shillong, Meghalaya, 793022, India (Ref: SEED/TIDE/007/2013/G) |
| Kim & Lee., 2016 | Randomized controlled trial | EEG recording (SMR and M-beta) | NI | n= 9 – received FES | EEG (Sensorimotor rhythm (SMR, 12–15 Hz) and mid-beta (M-beta, 15–20 Hz) waves were evaluated.) | NI |
| Larina et al., 2020 | Non-randomized trial | EEG recording | NI | NA | EEG | NI |
| Legarda et al., 2011 | Case report | TOVA, QEEG | NI | NA | Symptom profiles, TOVA, QEEG, and other observer evaluations. | No external funding |
| Neuper et al., 2003 | Case report | EEG | NI | NA | EEG, cursor control, letter selection accuracy | Austrian Federal Ministry of Transport, Innovation and |
|  |  |  |  |  |  | Technology, project GZ140.587/2. |
| Taheriena et al., 2015 | Non-randomized trial | The first session (introductory lesson) | NI | NA | Skill level, control of the virtual cube, puzzle game completion. | NI |
| Korandeh et al., 2021 | Quasi experimental research (randomized) | First EEG recording and PBS | NI | received only standard therapy | PBS | NI |
| Yu et al., 2012 | Randomized controlled trial | EEG, LOTCA, SEF. | NI | n=14 - no NFB | LOTCA and SEF | NI |
| Larina et al., 2019 | Randomized controlled trial | Spasticity measurements | NI | n=20 | MFS, MAS, MTS, MRC-SS | NI |
| Taherian et al., 2016 | Case report | Introductory session - motor imagery | Yes - 6 weeks post training | NA | Performance on motor imagery task. | NI |
| Xie et al., 2021 | Non-randomized trial | First training session | NI | NA | Changes in rPSD | Key Research and Development Program of Guangdong Province, China (#2018B030339001), in part by the National Natural Science Foundation of China (#81330032, #81771925, #61961160705, #U19A2082, and #61901077), in part by the National Key Research and Development Plan of China (#2017YFB1002501). |
| Korsunskaya et al., 2020 | Randomized controlled trial | MACS, ABILHAND-Kids, MFS, Ashworth scale, and Schulte tables, Münsterberg test | NI | r-Received NIMK-EX only | MACS, ABILHAND-Kids, MFS, Ashworth scale, and Schulte tables, Münsterberg test | Federal Target Program of the Ministry of Science and Higher Education of the Russian Federation, project number RFMEFI60519X0186. |
| Pavlenko et al., 2023 | Non-randomized controlled trial | MAS, ABILHAND- Kids, Assessment of oral praxis and articulatory motility, pronunciation of sounds, making sentences based on | NI | n=20, only received complex resort treatment | MAS, ABILHAND- Kids, Assessment of oral praxis and articulatory motility, pronunciation of sounds, making sentences based on | NI |
|  |  | the pictures. |  |  | the pictures. |  |
| Pavlenko et al., 2024 | Non-randomized trial | EEG recording at the first session, The Barthel Index, ABILHAND-Kids, MAS, MACS | NI | NA | EEG, Barthel Index, ABILHAND-Kids, MAS, MACS | Russian Science Foundation and the Republic of Crimea Grant no. 22-15-20035, https://rscf.ru/en/project/22-15-20035/. |
| Chen et al., 2024 | Randomized controlled trial | CPT-II, TVPS-3, and CPRS | NI | n= 11, usual rehabilitation | EEG, CPT-II, TVPS-3 and CPRS. | NI |
| Pavlenko et al., 2024 | Non-randomized trial | EEG (first session) and Barthel Index | NI | NA | EEG and Barthel Index | NI |
| EEG: Electroencephalography, NI: Not Indicated, SRTT: Serial Reaction Time Task, NA: Not applicable MAS: Modified Ashworth Scale, VSMS: Vineland Social Maturity Scale, SMR: Sensorimotor Rhythm, M-beta:Mid-beta, FES: Functional Electrical Stimulation, TOVA: Test of Variables of Attention, QEEG: Quantitative Electroencephalography, PBS: Pediatric Balance Scale, LOTCA: Loewenstein Occupational Therapy Cognitive Assessment, SEF: Somatosensory Evoked Fields, MFS: Motor Function Scale, MTS : Muscle Tone Scale, MRC-SS: Medical Research Council Sum Score, rPSD: Relative Power Spectral Density, MACS: Manual Ability Classification System, TVPS-3: Test of Visual-Perceptual Skills, Third Edition, CPT-II: Conners' Continuous Performance Test II, CRPS: Complex Regional Pain Syndrome |  |  |  |  |  |  |

**Table 3.** Summary of studies - Intervention characteristics.

| Study | Intervention | Duration | Device/Technology | Electrodes of Interest (10-20 system) | Place of intervention | Interventionist | Type of feedback | Frequency of Interest |
| --- | --- | --- | --- | --- | --- | --- | --- | --- |
| Alves-Pinto et al., 2017 | NF intervention - To find a mental condition that made video to be played. | 2 experimental sessions one over 30 mins. | EEG. Mindfield® Neuromaster 2-channel EEG system for neurofeedback (Mindfield® Biosystems Ltd., Germany). | 2 electrodes - C3 and C4 | Office room | Experimenter | Visual | Alpha power (8-13 Hz) |
| Filho et al., 2017 | NF intervention | Twenty sessions, three times per week, with each session lasting 45 minutes. | Pocket Neurobics Q-wiz EEG amplifier, Bio explorer software and TQ-7 system | 2 electrodes – C3 and C4 | Clinic | NI | NI | Sensorimotor rhythm (7.5-10.5 Hz) and inhibiting high beta waves (19-38 Hz) |
| Gupta et al., 2020 | NF intervention | 30 mins daily for 20 days | Bio Trace software for Nexus- 10 (through technology Inc Russia). | 8 electrodes – F3-F4, C3-C4, P3-P4, O3-O4 | NI | NI | NI | Alpha |
| Kim & Lee., 2016 | NF intervention/ BCI | 6 weeks, 5 times per week, 30 mins for a session | EEG (Brainwave-sensing headset, Neurosky, 2011), BCI-FES (EMG FES 1000-Walking Man II, Cybermedic Inc., Korea, 2009). | 2 electrodes - Fp1 and Fp2 | Rehabilitation hospital | NI | Kinaesthetic - FES | Sensorimotor rhythm (12–15 Hz) and mid-beta (15–20 Hz). |
| Larina et al., 2020 | NF intervention - motor imagery | 10 sessions, 3 trials each 8 minutes | EEG Neurovisor BMM electroencephalo | 21 electrodes – Fpz, Fp1, Fp2, Fz, F1, F2, F3, | Clinical Sanatorium | NI | Visual and kinaesthetic | Alpha |
|  |  |  | graph (Medical Computer Systems; Russia), Robotic hand (Exohand 2) | F4, Cz, C1, C2, C3, C4, Pz, P1, P2, P3, P4, Oz, O1, O2 |  |  |  |  |
| Legarda et al., 2011 | NF intervention | 20 separate 30 mins sessions over 4-8 weeks. | QEEG, Cygnet1 NF software integrated with Somatic Vision video game | 6 electrodes – T3-T4, C3-C4, T4-P4, T4-Fp2 | NI | NI | Visual | Infraslow oscillations. |
| Neuper et al., 2003 | NF intervention/BCI - motor imagery | Twice a week, 3-5 sessions lasting from 1.5 to 2 hours in total. | EEG, Windows with a 4-channel EEG amplifier | 4 electrodes | Rehabilitation center | NI | Visual and auditory | Upper beta band (20 – 30 Hz) |
| Taheriena et al., 2015 | NF intervention/BCI - motor imagery | 5-7 sessions. 3 weeks. 30 mins per session. | EEG (Emotiv Epoc), Emotiv's software | 2 electrodes – C3 and C4 | Special education school or in the participant's home. | NI | Visual | NI |
| Korandeh et al., 2021 | NF intervention | Twice weekly, each session lasted for half an hour for 5 weeks. | Procomp machine 2 made by Thought Company in Canada | 2 electrodes – O1 and O2 | NI | NI | Visual | Theta (4-7 Hz), beta (15-18 Hz) |
| Yu et al., 2012 | NF intervention | Twice a week for 30 minutes over a period of 6 weeks. | QEEG-8, NeuroComp System (Neurocybernetics Company, Missouri, America). | NI | Hospital | NI | Visual and auditory | SMR (12-15 Hz) or mid-Beta (15-18). |
| Larina et al., 2019 | NF intervention-motor imagery | 10 sessions, each with 3 sessions of 10 minutes per day | Exokist-2, Exohand-2 | NI | NI | NI | Visual and kinesthetic | NI |
| Taherian et al., 2016 | NF intervention/BCI - motor imagery | 4-week period, with three 30 min training per week. | Emotiv EPOC | 14 electrodes – AF3, AF4, F7, F3, FC5, T7, P3, P7, O1, O2, P8, P4, T8, FC6, F4, and F8 | School | NI | Visual | NI |
| Xie et al., 2021 | NF intervention/BCI - Motor imagery | Short term - 12 days 40 trial each day each 18 seconds<br>long term - 60 days, 40 trial day each 18 seconds | EEG (BrainMaster, Inc., Shenzhen, China), Symptom amplifier and 16 Ag/AgCl electrodes (BrainMaster, Inc., Shenzhen, China) | 16 electrodes – Cp1, Cp2, C3, C4, Po3, Po4, P3, P4, O1, O2, Cp3, Cp4, T3, T4, T5, and T6 | NI | Therapist | Visual | SMR (8-15 Hz) |
| Korsunskaya et al., 2020 | NF intervention/BCI - motor imagery | 10±2 days, lasting from 4 to 20 minutes | EEG (MKS-KEP), Exohand | NI | Medical center | NI | Kinesthetic | NI |
| Pavlenko et al., 2023 | NF intervention/BCI - motor imagery | 10 sessions of, 6 days in a week, three takes per session of 8 min with a rest break of at least 5 min | Exohand-2 | NI | Rehabilitation center | NI | Visual and kinesthetic | NI |
| Pavlenko et al., 2024 | NF intervention/BCI - motor imagery | 10 sessions, every other day, three takes per session of 8 min with a rest break of at least 5 min | EEG Neurovizor BMM-52 (Medical Computer Systems; Russia), Exohand-2 | 15 electrodes – F3, Fz, F4, FC3, FCz, FC4, C3, Cz, C4, CP3, CPz, CP4, P3, Pz, and P4 | NI | NI | Visual and kinesthetic | Mu (8-13 Hz) and Beta (15-25 Hz) |
| Chen et al., 2024 | NF intervention-game | 10-week, twice per week, and approximately 1 | EEG (Neuron Spectrum-2, Neurosoft, | 16 electrodes recording – training at C3-F3 | Laboratory room | Occupational therapist | Visual | Theta/Beta ratio |
|  |  | hour per time | Ivanovo) |  |  |  |  |  |
| Pavlenko et al., 2024 | NF intervention/BCI - motor imagery | 10 sessions, 6 days in a week, three takes per session, each take 8 minutes long, at least 5 minutes of rest between the takes. | EEG (BMM-52, (NVX-52 amplifier, Moscow,Zelenograd, MKS), Exokist-2 (Exoplast, Russia; Reg. Cert. RZN 2018/7681) | 10 electrode pairs – F3–C3, FS3–C3, C3–CP3, C3–P3, F3–P3, F4–C4, FS4–C4, C4–CP4, C4–P4, F4–P4, F3–F4, FS3–FS4, C3–C4, CP3–CP4, P3–P4 | Health center | NI | Visual and kinesthetic | Theta (4–8 Hz), Alpha (8–13 Hz), Beta1 (14–20 Hz), and Beta2 (20–30 Hz) |
| NF: Neurofeedback, BCI: Brain-Computer Interface, EEG: Electroencephalography, FES: Functional Electrical Stimulation, EMG: Electromyography, QEEG: Quantitative Electroencephalography, NI: Not Indicated, Hz: Hertz, SMR: Sensorimotor Rhythm, Mu: Mu rhythm |  |  |  |  |  |  |  |  |

### Outcome measurements

The outcome measurements of each study are reported in Table 2. These outcome measurements varied across studies and included assessment of brain activity, motor functions, cognitive functions, and task performance. Brain activity was assessed using EEG, focusing on changes measured before and after the intervention. Motor functions were assessed using the Modified Ashworth Scale (MAS),^65^ ABILHAND-Kids,^66^ Barthel Index,^67^ Modified Frenchay Scale (MFS),^68^ Modified Tardieu Scale (MTS),^69^ Medical Research Council Weakness Scale (MRC-SS),^70^ and Manual Ability Classification System (MACS).^71^ Cognitive functions were measured using Vineland Social Maturity Scale (VSMS),^72^ the Test of Variables of Attention (TOVA),^73^ and Loewenstein Occupational Therapy Cognitive Assessment (LOTCA).^74^ Task performance was assessed using matrices such as cursor control and/or puzzle game completion.

### Study quality appraisal and risk of bias assessment

The ROBINS-I V2 assessment is shown in Appendix S3, the RoB 2 in Appendix S4 and the SCD risk of bias tool in Appendix S5. The Downs and Black checklist is reported in Appendix S6, and SCED Scale is in Appendix S7.

The quality of evidence (SCED Scale) of the SCED studies ranged from 3/10 to 9/10, indicating a range from weak to high quality of evidence. Legarda et al.^63^ showed weak quality of evidence with the lowest scores in target behaviours, design, inter-rater reliability, independence of assessors, statistical analysis, and replication. Filho et al.,^64^ and Neuper et al.,^17^ showed moderate quality of evidence with the lowest scores in independence of assessors, replication and generalisation. Taherian et al.,^18^ showed high quality of evidence with the lowest scores in independence of assessors and statistical analysis. All SCED studies scored the maximum in baseline, sampling behaviour during treatment, and raw data recorded, and zero in independence of assessors. Based on the SCD risk of bias, the lowest risk of bias in all included studies was seen in participant selection and selective outcome reporting, while the highest risk of bias was seen in the sequence generation, blinding of participants and outcome assessors.

The quality of evidence (Downs and Black checklist) of the RCTs and non-RCTs ranged from 6/32 to 21/32, indicating poor to fair quality of evidence. Taherian et al.,^58^ showed the lowest quality of evidence with the lowest scores in external validity, internal validity, selection bias, and power. Kim and Lee^50^ showed the highest quality of evidence with the lowest scores for external validity. Based on the ROBINS-I V2, Xie et al.,^59^ had a critical risk of bias, whereas Kim and Lee^50^ (as assessed using the RoB 2) had a high risk of bias.

## Synthesis of results

### Brain outcomes

Out of the 18 included records, 13 studies^17, 18, 49–51, 53, 56, 57, 59, 61–64^ reported brain activity as an outcome measure, assessed using EEG before and after the intervention. These studies employed various EEG configurations, differing in the number and location of electrodes. Changes in brain activity were examined both within groups (pre-vs. post-intervention) and between groups (NF intervention vs. control conditions). Aiming at the inhibition of the high-beta frequency with NF training, Filho et al.,^64^ reported an inhibition in high-beta only in the C4 electrode site, but not in C3. Gupta et al.,^49^ found that although NF training had beneficial effects on brain activity, repetitive transcranial magnetic stimulation (r-TMS) was more effective in regulating neural activity. Kim and Lee^50^ compared BCI-based NF intervention with functional electrical stimulation (FES) alone, reporting that FES did not produce significant changes across electrodes, whereas BCI-based NF led to greater improvements. Larina et al.,^57^ demonstrated that BCI-based NF training increased neocortical activity and highlighted the influence of hemiparesis side on intervention outcomes. Neuper et al.,^17^ reported two distinct EEG patterns following BCI-based NF training. Yu et al.,^51^ measured changes in Spectral Edge Frequency (SEF), finding that while SEF 95% remained unchanged between NF and control groups, both groups showed an increase in SEF 50%. Xie et al.,^59^ showed that BCI-based NF training enhanced neural activity related to motor processing, with more pronounced effects in participants receiving ten days of therapy. Chen et al.,^53^ recorded a decrease in the theta/beta ratio following NF training, with improvements becoming more evident from early to middle (sessions 8–13) and late (sessions 14–19) phases of the intervention. Results are reported in Table 4.

**Table 4.**
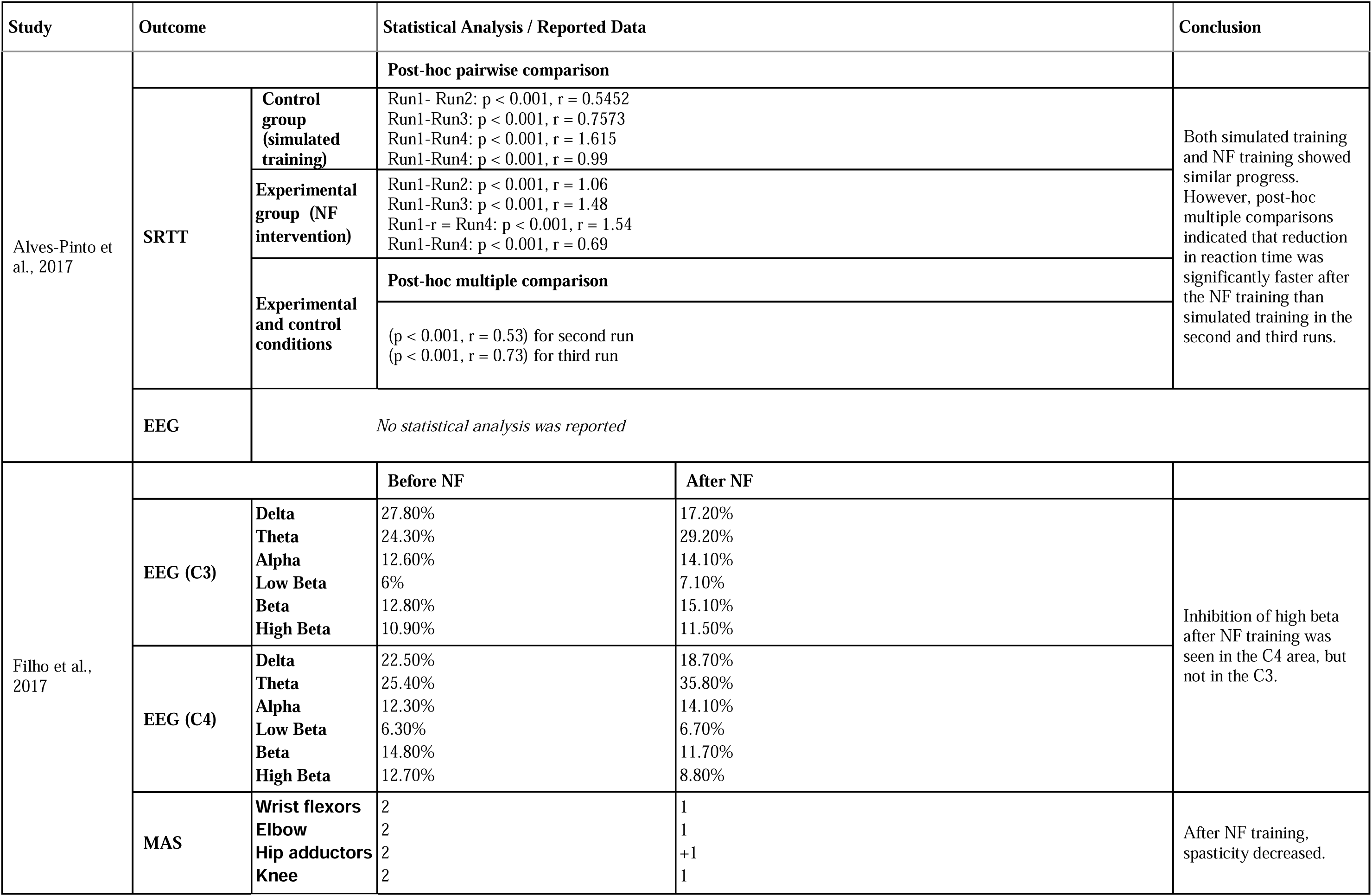
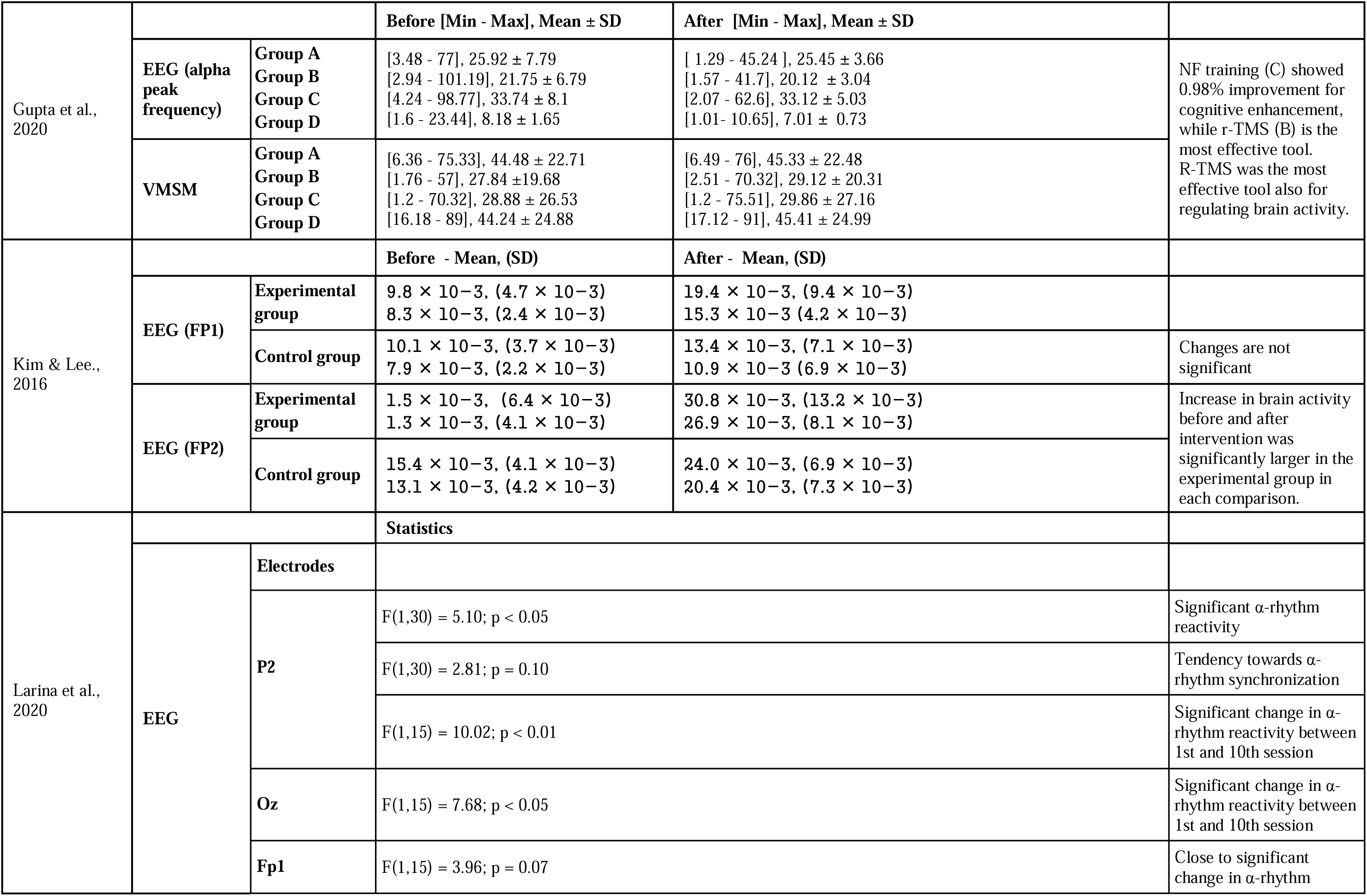
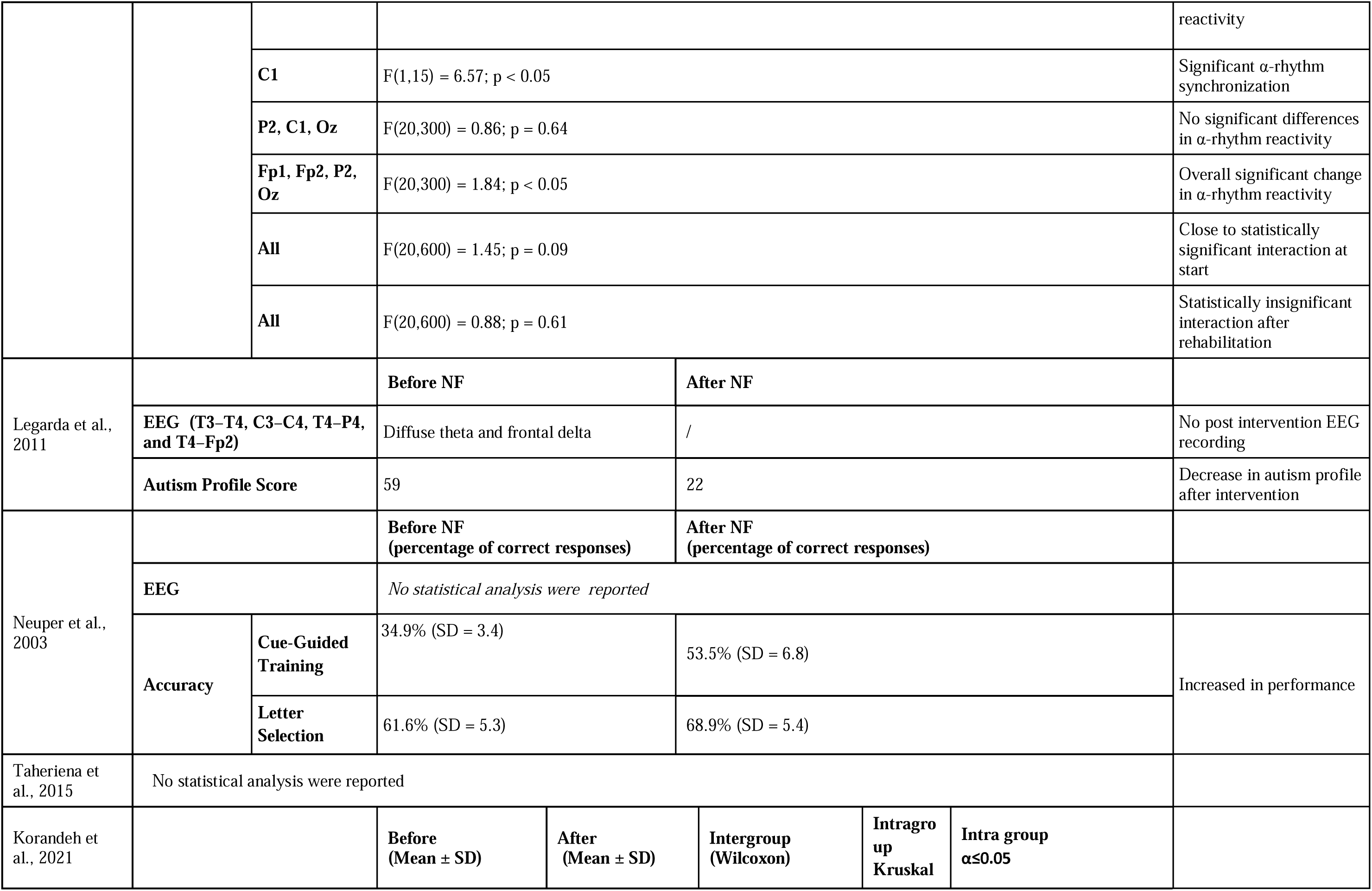
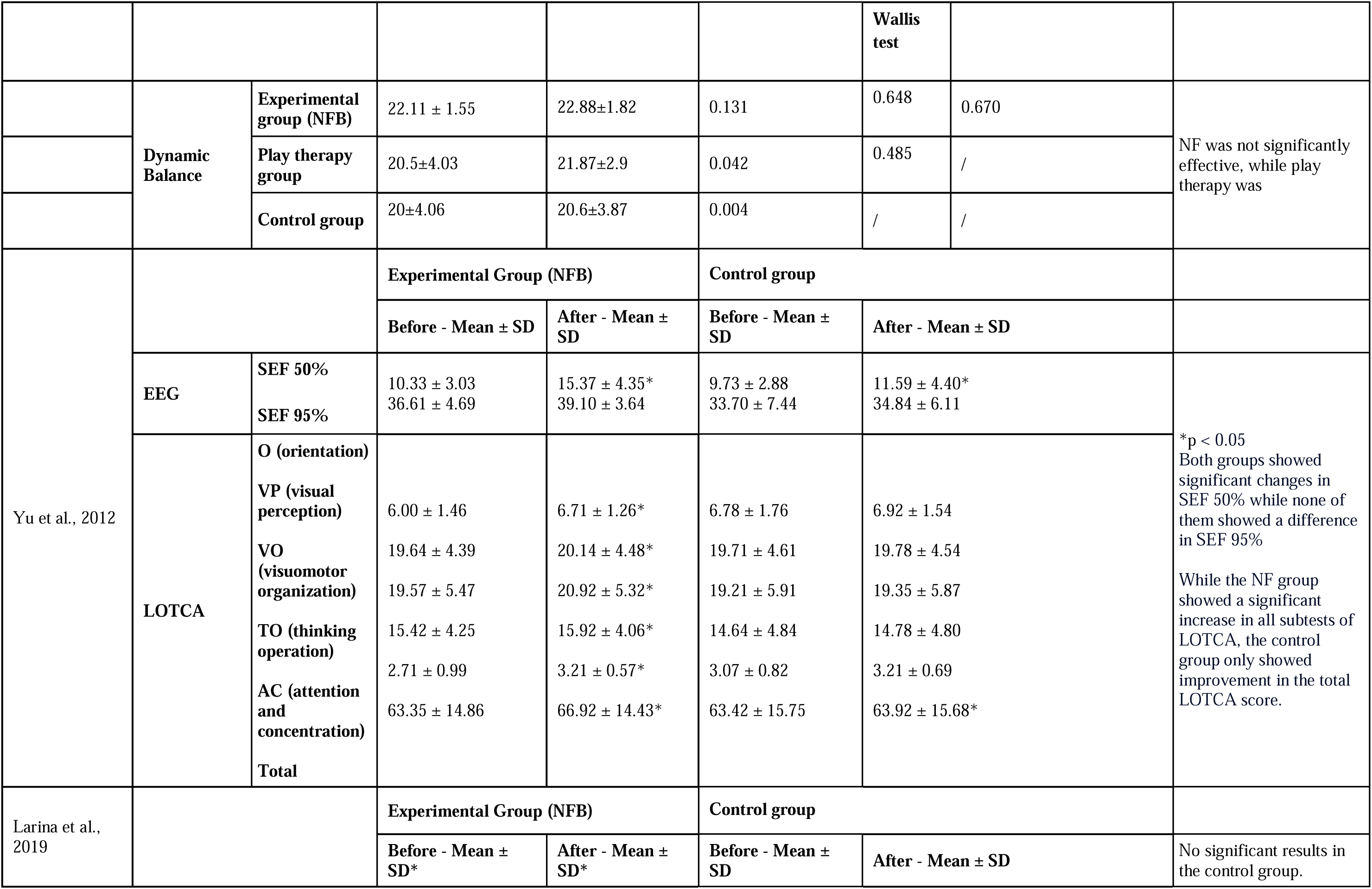
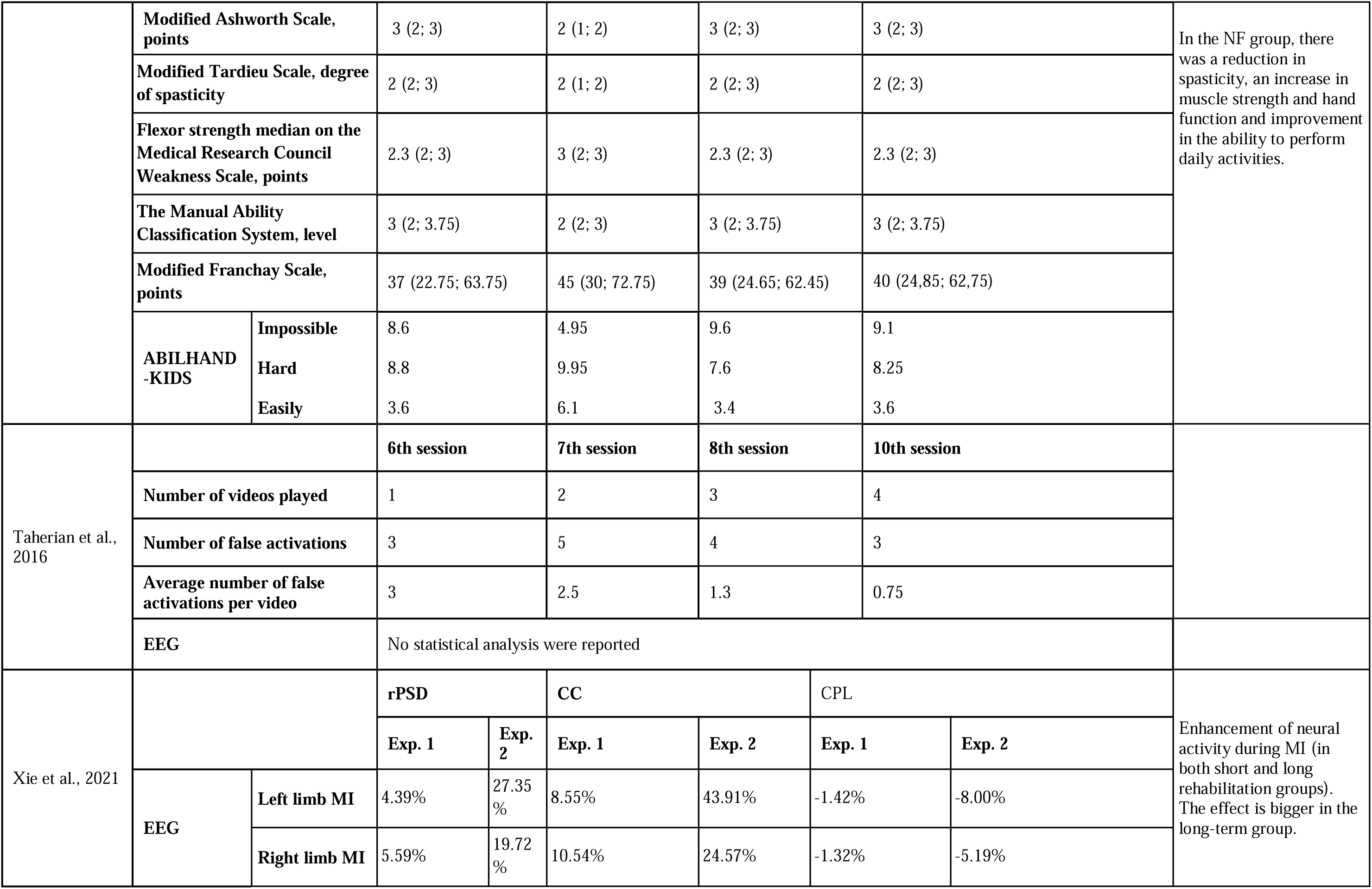
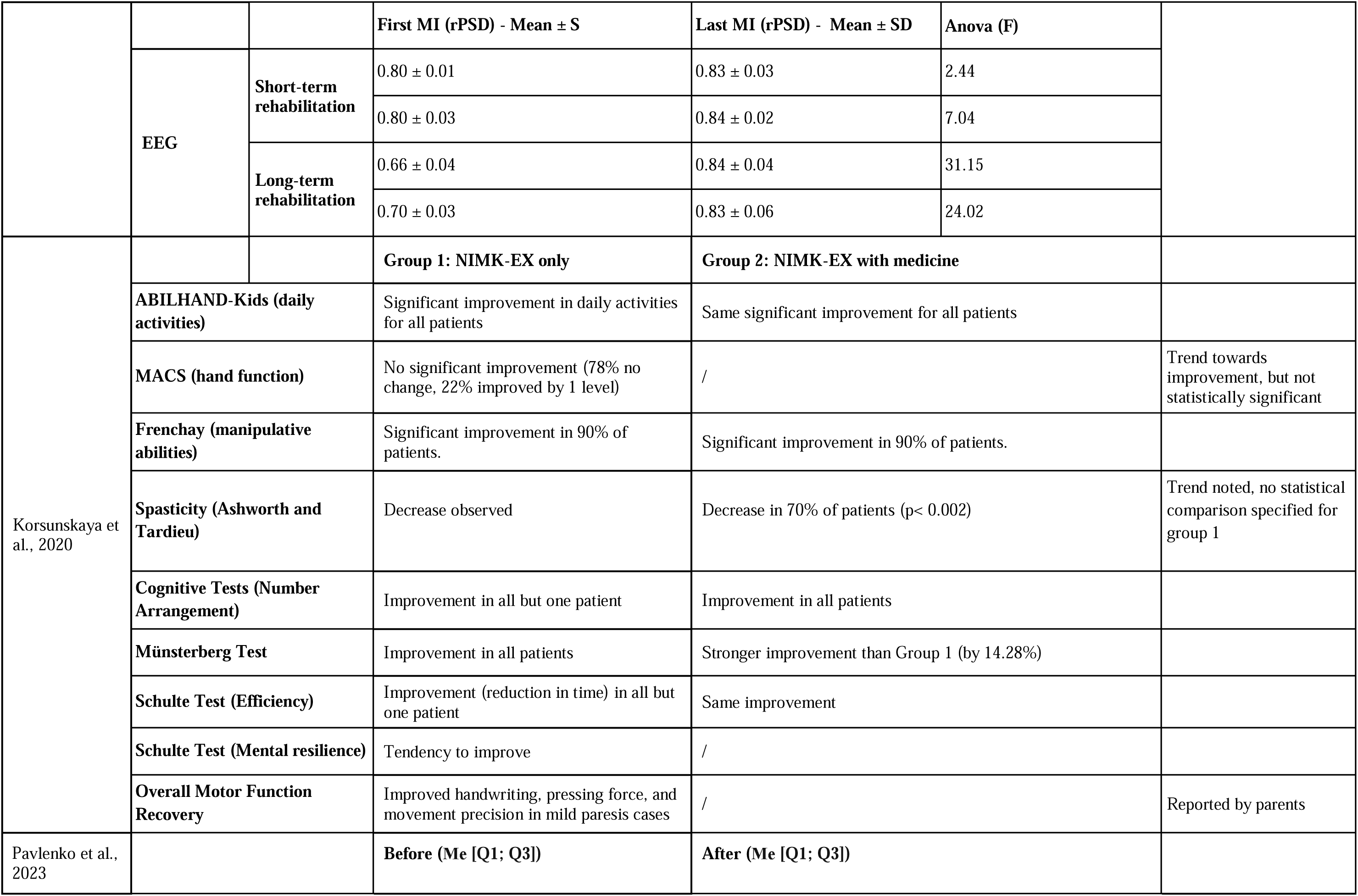
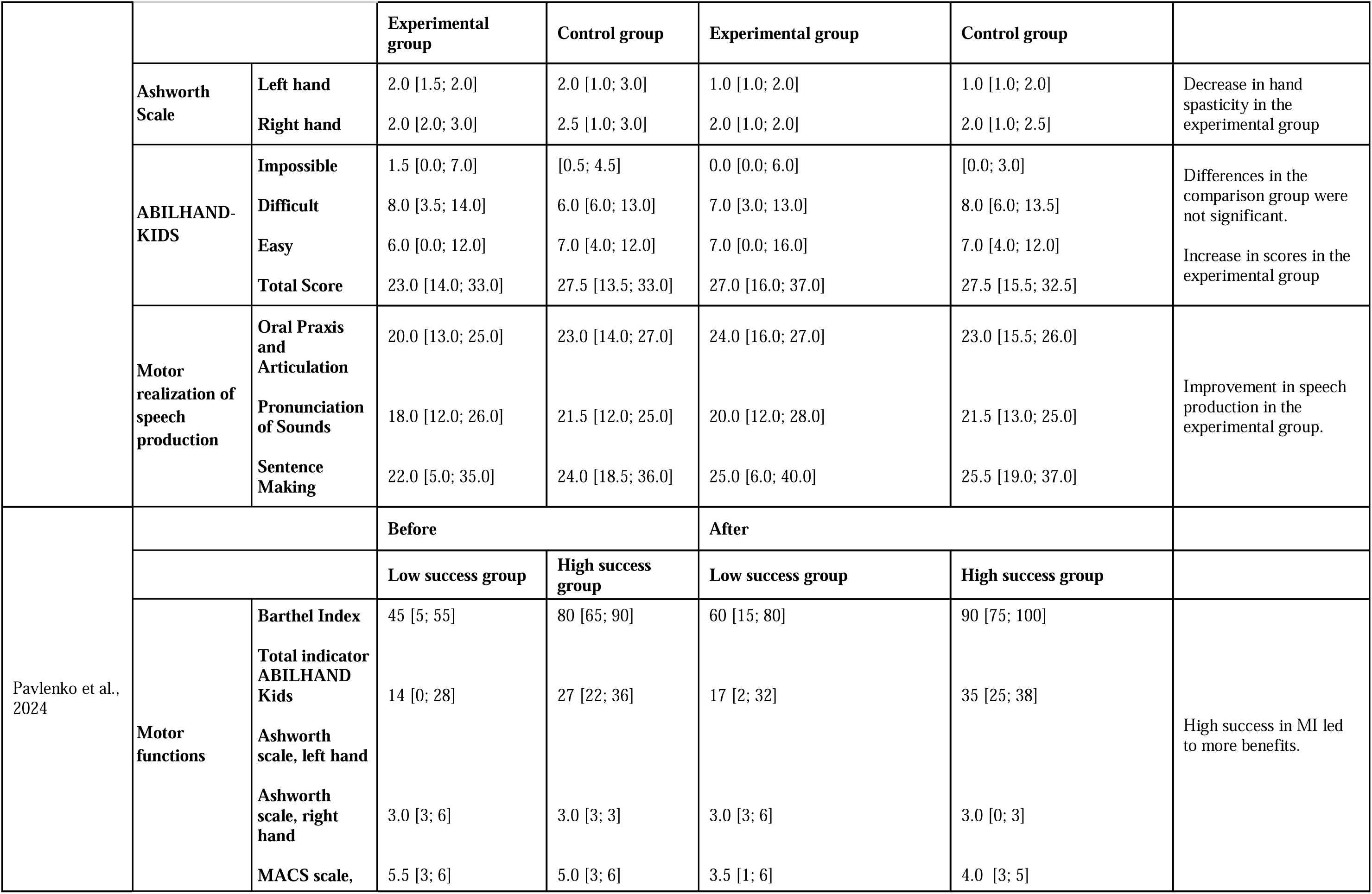
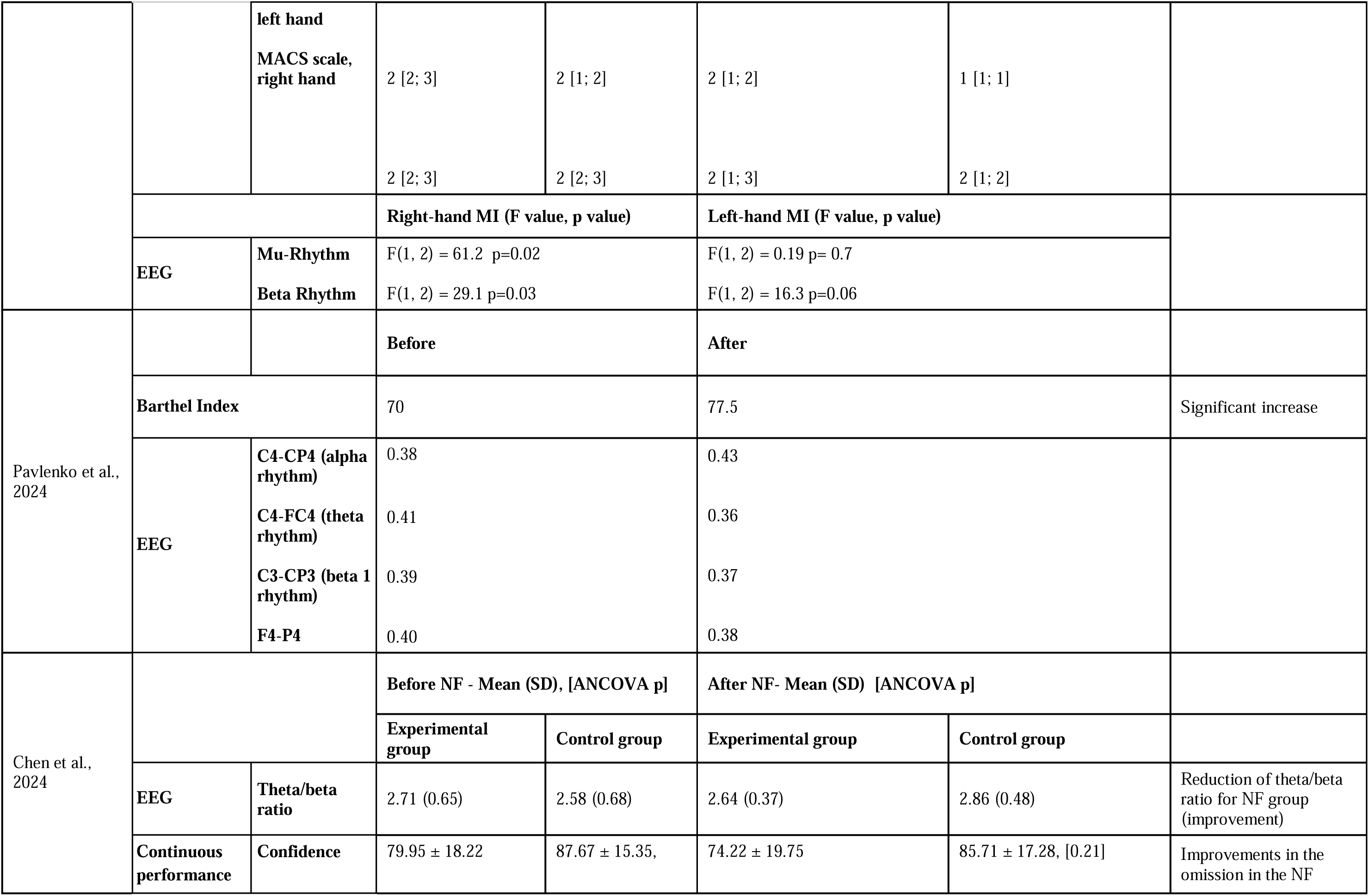
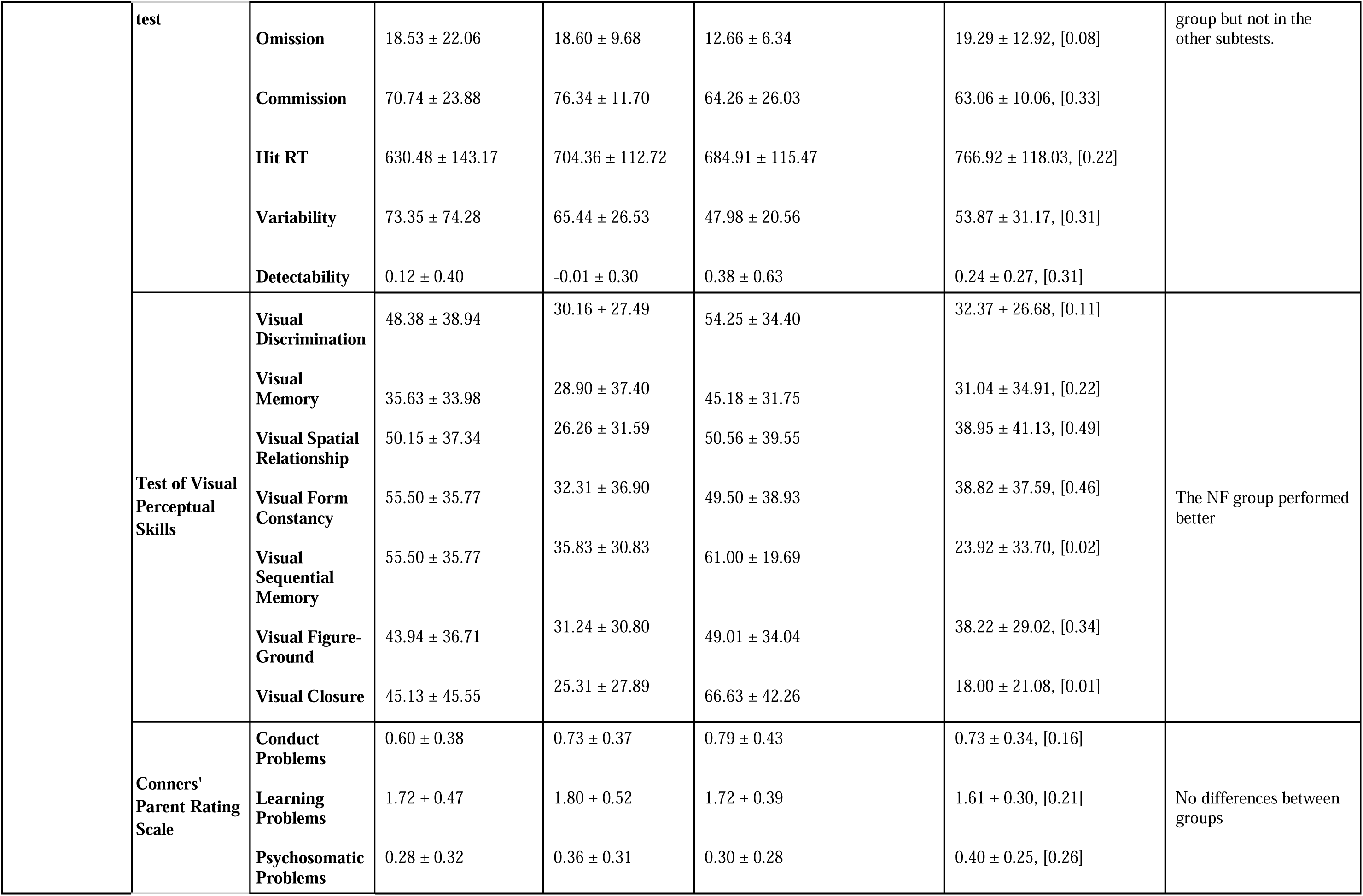
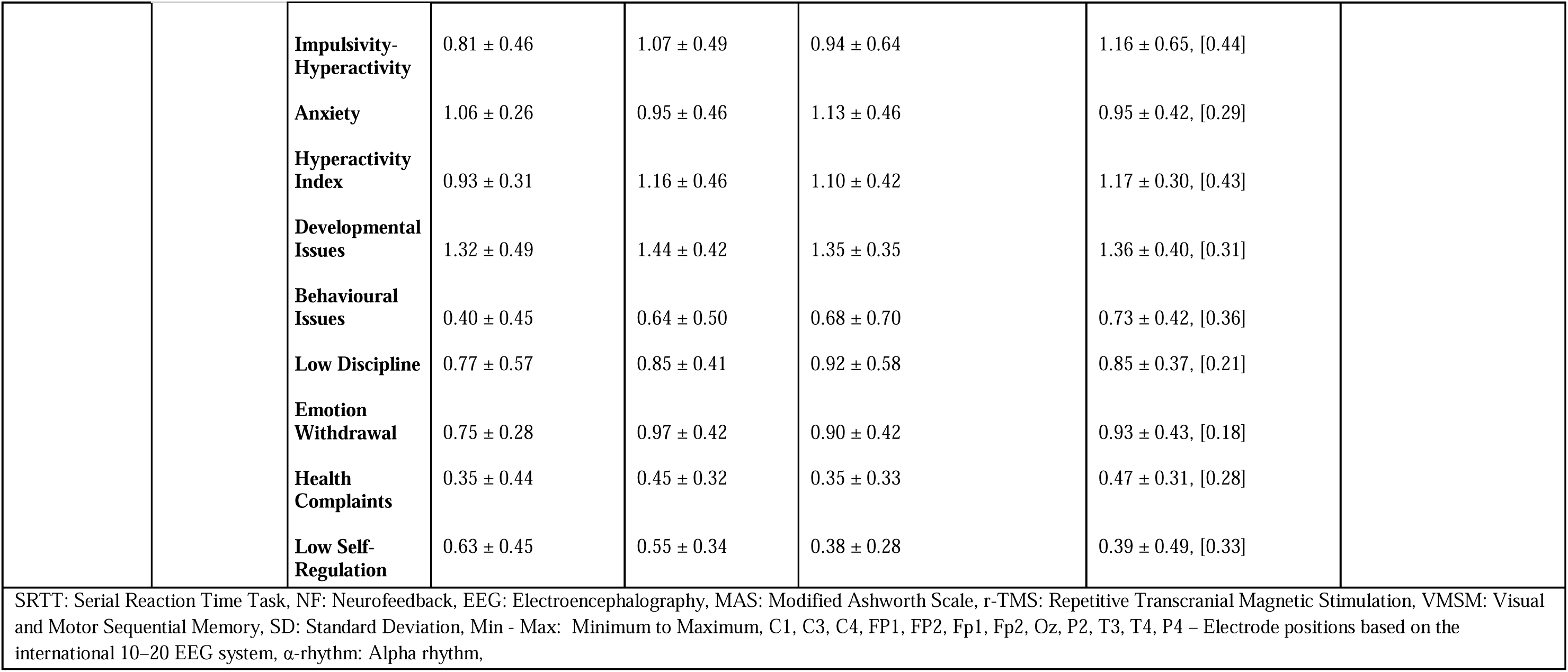
Outcome measures and results.

### Motor outcomes

Out of 18 studies, six included motor assessments as outcome measurements.^52, 54, 55, 60, 62, 64^ Filho et al.,^64^ showed decreased spasticity after NF training targeting the C3 and C4 electrodes, using an NF paradigm to reinforce SMR (7.5 - 10.5 Hz) and inhibit high beta (19 - 38 Hz). Korandeh et al.,^54^ showed that NF intervention was not significantly effective in improving dynamic balance, while play therapy led to significant improvements in dynamic balance. Larina et al.,^55^ demonstrated that NF intervention was effective in the reduction of spasticity, an increase in muscle strength and hand function and improved the ability to perform daily activities, while standard therapy did not lead to any significant improvements. Pavlenko et al.,^60^ showed that BCI-based NF interventions have a positive effect on decreasing spasticity as well as standard resort treatment (control group). Pavlenko et al.,^60^ showed that an increase in speech production was only seen in the experimental group, as well as an increase in daily activities reported by parents. Additionally, they demonstrated that high success in MI results in receiving more benefits from BCI-based NF interventions.^62^ Results are reported in Table 4.

### Cognitive outcomes

Out of 18 included records, five reported cognitive assessments as outcome measurements. ^17, 49, 51, 53, 56^ Alves-Pinto et al.,^56^ found that both simulated and actual NF training had similar effects on reaction time, although the reduction was more pronounced in the actual NF training group. Gupta et al.,^49^ showed that while NF training yielded significant positive effects, it was not the most effective method for cognitive enhancement when compared to repetitive transcranial magnetic stimulation (r-TMS), which demonstrated superior outcomes. Neuper et al.,^17^ reported improvements in letter selection ability, a skill that may support communication needs in individuals with CP. Yu et al.,^51^ observed that NF intervention led to improvements across multiple cognitive domains, including orientation, visual perception, visuomotor organisation, thinking operations, attention, and concentration, as measured by the LOTCA test. In contrast, the control group showed significant improvement only in the total LOTCA score. Lastly, Chen et al.,^53^ found that NF training improved visual-spatial memory abilities; however, no significant differences were observed between the NF and control groups in behavioural outcomes as rated by parents. Results are reported in Table 4.

### BCI-based neurofeedback

Out of 18 studies, nine used BCI-based NF.^17, 18, 50, 52, 58–62^ Several studies incorporated additional external devices, such as the Exohand-2 robotic hand or FES, to deliver feedback. All studies using BCI-based NF reported significant improvements, except one that yielded inconclusive results. Reported neural changes following BCI-based NF included significant modulation of SMR and M-beta frequency bands,^50^ SMR and mu rhythm,^62^ beta and alpha frequency bands,^17^ increased relative power spectral density (rPSD) during MI,^59^ and increased coherence in the alpha frequency band, the latter positively correlating with motor function improvements.^61^ Taherian et al.^18^ reported that enhanced control over brain activity led to improved use of an augmentative and alternative communication device, with effects persisting six weeks post-intervention. In another study by the same authors,^58^ inconclusive findings regarding the ability to control neural activity after training were reported, despite observing learning effects during the intervention. Beyond changes in brain activity, studies also showed improvements in motor function,^52, 60, 61^ everyday activity management and speech production,^60^ as well as cognitive outcomes, including enhanced attention.^52^ Results are reported in Table 4.

## DISCUSSION

This systematic review synthesises current evidence on the effects of NF interventions in individuals with CP. The included studies suggest that NF may have beneficial effects on brain function, motor performance, and cognitive outcomes. However, the overall strength of the evidence is limited by methodological variability across studies. Factors such as the type of feedback used, the targeted frequency bands, the duration and intensity of the intervention, and the heterogeneity in clinical characteristics of participants with CP may influence the efficacy of NF interventions.

### Neurofeedback intervention protocol

This systematic review highlights substantial heterogeneity in EEG-based NF protocols used in individuals with CP. Although all included studies employed EEG to deliver NF, there was considerable variation in intervention protocols, including the type of feedback, frequency bands targeted, number and placement of electrodes, and the equipment used. Such variability poses challenges for generalising the effects of NF interventions across studies.

Notably, even among studies targeting similar outcomes, such as motor imagery (MI) training, differences in electrode configuration were evident. For instance, one study used only two electrodes, while another employed fourteen, despite both aiming to modulate similar brain functions. This inconsistency in electrode number and placement has also been observed in NF research involving healthy adults, as reported by Rogala et al.^75^ Electrode selection is influenced by several factors, including individual anatomical differences, the need to measure average activity across regions, and the specific NF approach employed. Dense electrode arrays may help account for individual variability in EEG mapping due to anatomical differences,^76^ while selecting multiple neighbouring electrodes can improve the accuracy of average activity measurements.^77^ Furthermore, a higher number of electrodes enhances localisation precision, which is particularly important for source-level NF training.^78^ Given these considerations, individualised electrode localisation may offer the most effective approach for optimising NF outcomes.

The majority of studies included in this review employed visual feedback as the primary method during NF training, while some combined visual feedback with kinaesthetic modalities. In studies utilising BCI-based NF, kinaesthetic feedback was delivered through external devices such as hand exoskeletons, FES, or powered mobility systems. The modality of feedback, whether visual, kinaesthetic, auditory, or a combination, plays a critical role in determining the efficacy of NF interventions. For MI training, which was a common approach among the included studies, evidence from healthy participants suggests that anatomically realistic visual feedback (e.g., animations aligned with the participant’s actual hand position) yields greater benefits than non-specific visual representations.^79^ Moreover, combining visual feedback with realistic kinaesthetic input via external devices has been shown to enhance the effectiveness of NF training beyond visual feedback alone.^80^ These findings underscore the importance of selecting feedback modalities that are both functionally relevant and tailored to the user’s capabilities and therapeutic goals.^22^

### Efficacy of neurofeedback intervention

This review generally indicates positive effects of NF in individuals with CP, although the results remain open to interpretation. For instance, a placebo-controlled study in which the control group received simulated NF showed similar cognitive improvements to the experimental group receiving actual NF training.^56^ Additionally, another study reported brain-related improvements in a control group undergoing traditional therapy.^51^ These findings suggest that the efficacy of NF in CP warrants further investigation.

NF interventions have been shown to improve cognitive outcomes in children with ASD,^26^ brain outcomes in ASD,^81^ and cognitive outcomes in ADHD.^82^ However, a separate review did not support the positive effects of NF in children with ASD.^25^ This discrepancy may be attributed to placebo effects or variations in intervention protocols. The potential placebo effect of NF has been discussed in other studies.^83, 84^

The efficacy of NF appears to depend significantly on the intervention protocol. In this review, improvements in brain activity were more pronounced following long-term therapy sessions, suggesting that the duration of intervention plays a critical role in achieving beneficial outcomes. This may be attributed to the brain’s learning capacity (i.e. neuroplasticity), which underlies the lasting effects of NF training.^12^ A meta-analysis has shown that NF activates specific brain regions associated with neuroplasticity.^85^ Individual differences, such as learning capacity, may also influence how effectively participants benefit from NF.^86^ One study included in this review compared children with higher and lower success in MI, reporting that the group with higher MI success demonstrated greater cognitive improvements following NF. The authors suggested that better MI performance enhanced the classifier’s ability to recognise EEG patterns, potentially accelerating the learning process from feedback. Consequently, it has been proposed that the frequency and number of NF sessions should be personalised based on the participant’s learning curve.^77^ Beyond session frequency, individual differences in oscillatory brain activity should also be considered when tailoring NF protocols to maximise therapeutic outcomes.^87^

### Neurofeedback versus Repetitive Transcranial Magnetic Stimulation

While NF has demonstrated beneficial effects on modulating brain activity and enhancing cognitive outcomes, comparative studies suggest that r-TMS may be more effective in regulating brain activity. One study directly comparing r-TMS and NF found that r-TMS produced stronger effects on brain regulation. This aligns with findings from research in Parkinson’s disease, where a combination of EEG-NF and r-TMS yielded the most significant improvements in both brain and motor outcomes.^88^ However, r-TMS alone was more effective than NF alone, and similarly, in terms of cognitive outcomes, r-TMS outperformed NF training.^88^ A possible explanation for these differences lies in the mechanisms of action: r-TMS delivers direct, exogenous stimulation to the brain, which may result in faster and more robust effects. In contrast, NF is an endogenous, user-dependent method that relies heavily on the participant’s engagement and active participation. This dependency can pose challenges, particularly in paediatric populations such as children with CP, where maintaining consistent engagement may be difficult.

### Neurofeedback effectiveness based on the ICF domains

This review highlights the positive effects of NF across several domains of the ICF, particularly in body functions and structures, as well as activity and participation. NF training has shown beneficial effects on brain outcomes; however, these are typically phasic, event-related changes in oscillatory activity triggered by feedback.^89^ Due to the lack of longitudinal follow-up, it remains uncertain whether these changes translate into tonic, lasting alterations that impact everyday functioning. Positive effects have been observed in motor and cognitive outcomes, such as reduced spasticity, increased muscle strength, improved hand function, and improved executive functions and higher-order cognitive abilities. A possible explanation is that training modulation of SMR may underpin both motor and cognitive improvements, as highlighted in a recent systematic review and meta-analysis showing clinical benefits of SMR-based NF across these domains.^90^ Although these findings are not consistently supported by follow-up assessments, they provide promising avenues worth exploring in future longitudinal research with more robust methodology.

Benefits in the activity and participation domains were also identified, though fewer studies included these outcomes. The effects were less pronounced compared to brain, cognitive, and motor outcomes, possibly due to the challenge of capturing NF-related improvements in real-life contexts. Enhancements in speech production and letter selection, for example, could significantly empower children by boosting their confidence and enabling more meaningful interaction with their environment.

Contextual factors such as environmental and personal factors remain largely unexplored in NF research. Environmental factors such as family support, access to technology and inclusive settings may significantly influence the success of NF interventions.^91^ Similarly, personal factors like motivation, emotional factors, coping, and cognitive psychological factors are known to affect rehabilitation outcomes^92^ but they are not yet systematically studied in NF contexts. Future research should aim to systematically include and assess these contextual domains to better understand their role in shaping NF outcomes, particularly in paediatric populations with complex needs.

### BCI-based neurofeedback

This review highlights the promising potential of BCI-based NF in improving mobility and communication, both of which are essential aspects of functioning and fundamental rights. These technologies are particularly relevant for children with severe CP, who often experience significant limitations in voluntary motor control and coordination. BCI systems have been successfully used to support communication and mobility by enabling control of external devices such as wheelchairs^16^ and robotic hands.^93^ Studies show that when BCI is combined with assistive technologies, the outcomes are consistently more favourable, suggesting that integrating multiple modalities may enhance NF effectiveness by providing richer and more engaging feedback. Despite these encouraging results, research in paediatric populations remains limited.

Future studies should investigate the long-term effects and usability of BCI systems in children and develop age-appropriate interfaces that consider cognitive development, attention span, and emotional engagement. Addressing these gaps could help optimise BCI-based NF applications and ensure they are both effective and accessible for children with complex motor impairments.

### Limitations and directions for future research

This systematic review employed a comprehensive search strategy, including both English and non-English literature, and applied rigorous quality and risk of bias assessments to evaluate the included studies. Despite these strengths, several limitations must be addressed.

The overall methodological quality of the included studies ranged from weak to moderate, with a high risk of bias across most studies. Only one out of the 18 studies included a follow-up assessment, which significantly limits the ability to evaluate the sustained effects of NF interventions.^94, 95^ Follow-up data are essential for determining the long-term efficacy and reliability of NF in improving brain, motor, and cognitive outcomes in individuals with CP. In the absence of such data, the durability of intervention effects remains unknown.

Future research should not only prioritise well-designed RCTs with long-term follow-up but also address the lack of standardisation in NF protocols, including consistency in frequency bands, electrode placement, and session intensity. Beyond neural outcomes, trials should incorporate clinically meaningful measures such as quality of life, societal participation, and caregiver-reported outcomes to better capture real-world impact. Evaluating cost-effectiveness and accessibility is essential, particularly in CP, where families often face high care burdens and limited access to advanced rehabilitation technologies.^96^ Such efforts will help determine whether NF can become a sustainable intervention in routine clinical care.

Another methodological concern is the absence of power analyses in most studies, which hinders the interpretation of statistical and clinical significance. Although the Downs and Black checklist assigns the highest score for studies with more than eight participants, this criterion may be overly generous and insufficient for evaluating statistical power, especially in studies with small sample sizes.

Clinical variability also contributed to the complexity of this review. Participants differed in CP severity, comorbid conditions, age, and timing of outcome assessments. Implementing NF in children with CP may present practical challenges, including limited attention span, reduced motivation, fatigue, and the severity of associated movement disorders, all of which can affect training adherence and outcomes. To address these barriers, gamified approaches and VR-augmented NF may enhance engagement and motivation during sessions.^97^ This approach may be particularly valuable in children with severe CP, for whom conventional NF protocols may otherwise be difficult to tolerate. Participants across the included studies ranged from 4 to 47 years, covering both paediatric and adult populations. Most studies focused on children and adolescents, with only a few involving adults. No consistent age-related differences in NF effectiveness were reported, although developmental stage may influence engagement, learning capacity, and neural plasticity. These differences, along with variability in intervention protocols, introduced heterogeneity that was addressed through narrative synthesis rather than meta-analysis.

While these limitations highlight gaps in the current evidence base, the present review adhered to AMSTAR-2 methodological standards,^98, 99^ thereby ensuring good scholarship principles, transparency, and robustness in the synthesis.

## Conclusion

Despite methodological limitations and considerable heterogeneity in NF protocols across the included studies, this systematic review provides preliminary evidence that individuals with CP can learn how to regulate neural activity through NF. Findings support the potential of NF to improve brain function, motor performance, and cognitive outcomes in individuals with CP. By consolidating findings from diverse interventions and participant profiles, the review offers a foundational understanding of how NF may contribute to neurorehabilitation in this population. The integration of BCI and external devices (e.g. robotic hands, powered wheelchairs) appeared to further strengthen NF intervention effects. This is the first systematic synthesis focused specifically on NF applications in CP, highlighting both the promise of this emerging field and the need for more rigorous, standardised research. The insights gained from this review can inform the development of individualised NF protocols and guide future studies toward more robust designs, ultimately supporting clinicians and researchers in optimising therapeutic strategies for individuals with CP. Evaluating cost-effectiveness and the feasibility of clinical implementation is also essential for the translation of NF interventions into everyday rehabilitation settings.

## Supporting information

The three search strategies used in each database are shown in Appendix S1.

The list of excluded records, along with the reasons for exclusion, is provided in Appendix S2.

The ROBINS-I V2 assessment is shown in Appendix S3

the RoB 2 in Appendix S4

and the SCD risk of bias tool in Appendix S5.

The Downs and Black checklist is reported in Appendix S6,

and SCED Scale is in Appendix S7.

## Data Availability

All data produced in the present study are available upon reasonable request to the authors

## Acknowledgments

The authors have no interests that might be perceived as posing a conflict or bias. No funding or financial support was received for writing this systematic review.

