## Supplementary material for "Effectiveness of neurofeedback interventions in cerebral palsy: A systematic review": The three search strategies used in each database are shown in Appendix S1.

**Appendix S1. Search strategy**

**a) MEDLINE (Ovid)**

Two separate search strategies were used.

1- Advances search:

Per keyword:

1. neurofeedback
2. brain wave
3. eeg biofeedback
4. neurotherap*
5. cerebral palsy
6. 1 OR 2 OR 3 OR 4
7. 5 AND 6

2- Advanced Search

Per Keyword

1. brain-computer interface
2. brain computer interface
3. brain-machine interface
4. brain machine interface
5. cerebral palsy
6. 1 OR 2 OR 3 OR 4
7. 5 AND 6

**b) EMBASE**

Two separate search strategies were used.

1- Advanced search:

Per keyword:

1. #3 ‘neurofeedback’
2. #4 ‘eeg biofeedback’
3. #5 ‘neurotherap*’
4. #6 ‘brain wave’
5. #7 ‘cerebral palsy’
6. #8: #3 OR #4 OR #5 OR #6
7. #9: #8 AND #7

2- Advanced search

('brain computer interface'/exp OR 'brain computer interface' OR ('brain'/exp OR brain) AND (computer/exp OR computer) AND (interface/exp OR interface)) OR (brain computer AND interface) OR ('brain machine' AND interface) OR (brain AND machine AND interface)) AND cerebral AND palsy

**c) ProQuest**

Two separate search strategies were used.

1- Advanced search: Each combination of keywords using AND.

1. “cerebral palsy” AND “brain wave”
2. “cerebral palsy” AND “eeg biofeedback”
3. “cerebral palsy” AND neurofeedback
4. “cerebral palsy” AND neurotherap*

2- Advanced search: Each combination of keywords using AND.

1. “brain-computer interface” AND “cerebral palsy”
2. “brain machine interface” AND “cerebral palsy”
3. “brain-machine interface” AND “ cerebral palsy”
4. “brain computer interface” AND “cerebral palsy”

**d) Web of Science**

Two separate search strategies were used.

1. Advance Search: ((((ALL=(neurofeedback)) OR ALL=("brain wave")) OR ALL=("eeg biofeedback")) OR ALL=(neurotherap*)) AND ALL=("cerebral palsy")
2. Advance Search: ((((ALL=("brain-computer interface")) OR ALL=("brain computer interface")) OR ALL=("brain-machine interface")) OR ALL=("brain machine interface")) AND ALL=("cerebral palsy")

**e) PubMed**

Two separate search strategies were used

1. Advanced search (All fields): ("neurofeedback"[All Fields] OR "neurotherap*"[All Fields] OR "eeg biofeedback"[All Fields] OR "brain wave"[All Fields]) AND "cerebral palsy"[All Fields]
2. Advanced search (All fields): (((("brain-computer interface") OR ("brain computer interface")) OR ("brain-machine interface")) OR ("brain machine interface")) AND ("cerebral palsy")

**f) Scopus**

Two separate search strategies were used

1- Basic Keyword Search

1. ALL ( neurofeedback )
2. ALL ( "eeg biofeedback" )
3. ALL ( "brain wave" )
4. ALL ( neurotherap* )
5. ALL ( "cerebral palsy" )
6. ( ALL ( neurofeedback ) ) OR ( ALL ( "eeg biofeedback" ) ) OR ( ALL ( "brain wave" ) ) OR ( ALL ( neurotherap* ) )
7. ( ( ALL ( neurofeedback ) ) OR ( ALL ( "eeg biofeedback" ) ) OR ( ALL ( "brain wave" ) ) OR ( ALL ( neurotherap* ) ) ) AND ( ALL ( "cerebral palsy" ) )

2- Basic Keyword Search

1. ALL (“brain-computer interface”)
2. ALL (“brain computer interface”)
3. ALL (“brain-machine interface”)
4. ALL (“brain machine interface”)
5. ALL (“cerebral palsy”)
6. ( ALL ( "brain-computer interface" ) ) OR ( ALL ( "brain computer interface" ) ) OR ( ALL ( "brain-machine interface" ) ) OR ( ALL ( "brain machine interface" ) )
7. ( ( ALL ( "brain-computer interface" ) ) OR ( ALL ( "brain computer interface" ) ) OR ( ALL ( "brain-machine interface" ) ) OR ( ALL ( "brain machine interface" ) ) ) AND ( ALL ( "cerebral palsy" ) )

**g) CINAHL (EBSCO)**

Two separate search strategies were used

1. Advanced Search: (TX neurofeedback OR TX "eeg biofeedback" OR TX "brain wave" OR TX neurotherap*) AND TX "cerebral palsy"
2. (TX "brain-computer interface" OR TX "brain computer interface" OR TX "brain-machine interface" OR TX "brain machine interface") AND TX "cerebral palsy"

TX, all text

**h) Cochrane Library**

Two separate search strategies were used

1- Advanced Search

Search #1 neurofeedback AND “cerebral palsy”

Search #2 neurotherap* AND "cerebral palsy"

Search #3 "eeg biofeedback" AND "cerebral palsy”

Search #4 "brain wave" AND "cerebral palsy"

2- Advanced Search

“brain-computer interface” AND “cerebral palsy”

“brain computer interface” AND “cerebral palsy”

“brain-machine interface” AND “cerebral palsy”

“brain machine interface” AND “cerebral palsy”

**i) PsycInfo**

Two separate search strategies were used

1- Advanced search: (TX neurofeedback OR TX "eeg biofeedback" OR TX "brain wave" OR TX neurotherap*) AND TX "cerebral palsy"

2- Advanced search: (TX "brain-computer interface" OR TX "brain computer interface" OR TX "brain-machine interface" OR TX "brain machine interface") AND TX "cerebral palsy"

TX, all text
