## Supplementary material for "Effectiveness of neurofeedback interventions in cerebral palsy: A systematic review": The list of excluded records, along with the reasons for exclusion, is provided in Appendix S2.

| **Appendix S2: Excluded references in English and non-English language** | |
| --- | --- |
| ***English language references excluded after full-text review (n=76)*** | |
| **Study** | **Reason for Exclusion** |
| Filho et al., 2021^1^ | No NF intervention |
| Ayers, 2004^2^ | Opinion paper |
| Behboodi et al., 2023^3^ | No NF intervention |
| Behboodi et al., 2019^4^ | No NF intervention |
| Craig et al., 2002^5^ | No NF intervention/ No CP |
| Durham et al., 2004^6^ | No NF intervention |
| El-Shamy & El Kafy, 2021^7^ | No NF intervention |
| Wysoczanska & Skrzek, 2017^8^ | No NF intervention |
| Redstone, 2006^9^ | No NF intervention |
| Floreani et al., 2022^10^ | No NF intervention |
| Gad et al., 2021^11^ | No NF intervention |
| Girshin et al., 2023^12^ | No NF intervention |
| Gupta & Bhatia., 2018^13^ | No NF intervention |
| Gupta & Bhatia., 2021^14^ | Conference paper |
| Hammond, 2006^15^ | Opinion paper |
| Hastings et al., 2022^16^ | No NF intervention |
| Hosseini et al., 2023^17^ | No NF intervention |
| Kim et al., 2014^18^ | No NF intervention |
| Kim et al., 2015^19^ | No NF intervention |
| Klobucká et al., 2020^20^ | No NF intervention |
| Levac et al., 2018^21^ | No NF intervention |
| Lin & Shieh, 2014^22^ | No CP |
| Lins et al., 2019^23^ | No NF intervention |
| MacIntosh et al., 2020^24^ | No NF intervention |
| Rubi, 2007^25^ | Opinion paper |
| Mellinger et al., 2007^26^ | No NF intervention/ No CP |
| Myers & Young, 2012^27^ | Overview paper |
| Nam & Bahn, 2012^28^ | No NF intervention/ No CP |
| Parhizgar et al., 2012^29^ | No NF intervention/ No CP |
| Dahlstrand et al., 2023^30^ | No NF intervention |
| Rios et al., 2013^31^ | No NF intervention |
| Romero-Ayuso, 2020^32^ | No NF intervention/ No CP |
| Rose et al., 2017^33^ | Review |
| Sakamaki et al., 2022^34^ | No CP |
| Schertz et al., 2016^35^ | No NF intervention |
| Shafaei et al., 2022^36^ | No NF intervention |
| Sung et al., 2021^37^ | No NF intervention/ No CP |
| Syam et al., 2017^38^ | No NF intervention/ No CP |
| Vasilijevic & Miranda, 2023^39^ | No NF intervention/ No CP |
| Zahradka et al., 2021^40^ | No NF intervention |
| Ramakrishnan, 2024^41^ | No CP |
| An et al., 2024^42^ | No NF intervention |
| Jadavji et al., 2023^43^ | No NF intervention |
| Boulay et al., 2024^44^ | No NF intervention |
| Molina-Cantero et al., 2017^45^ | No NF intervention |
| Scherer et al., 2015^46^ | No NF intervention |
| Almeida et al., 2023^47^ | Conference paper |
| Augenstein et al., 2024^48^ | No NF intervention/ No CP |
| Bobrova et al., 2023^49^ | No NF intervention/ No CP |
| Cano-Izquierdo et al., 2023^50^ | No NF intervention/ No CP |
| Chen et al., 2024^51^ | No NF intervention/ No CP |
| Wu et al., 2023^52^ | No NF intervention/ No CP |
| Kelly et al., 2023^53^ | Conference paper |
| Ferracuti et al., 2023^54^ | No NF intervention/ No CP |
| Flux et al., 2023^55^ | No NF intervention |
| Hernandez-Mustieles et al., 2024^56^ | No NF intervention/ No CP |
| Chiang et al., 2023^57^ | No NF intervention/ No CP |
| Keough et al., 2024^58^ | No CP |
| Liu et al., 2024^59^ | No NF intervention/ No CP |
| Meng et al., 2024^60^ | Review |
| Miziara et al., 2024^61^ | No NF intervention |
| Moufassih et al., 2023^62^ | No NF intervention/ No CP |
| Raj & Mukul, 2023^63^ | Conference paper |
| Ranzani et al., 2024^64^ | No NF intervention/ No CP |
| Saltzman et al., 2024^65^ | No NF intervention/ No CP |
| Sawai et al., 2023^66^ | No CP |
| Siddiqui et al., 2023^67^ | No NF intervention/ No CP |
| Sreeja & Samanta, 2023^68^ | No NF intervention/ No CP |
| Tou et al., 2023^69^ | Not peer reviewed - preprint |
| Zhou et al., 2023^70^ | Review |
| Youssofzadeh et al., 2023^71^ | No NF intervention/ No CP |
| Wong et al., 2023^72^ | No NF intervention/ No CP |
| Wong et al., 2022^73^ | Perspective paper |
| Urrea & Agramonte, 2023^74^ | No NF intervention/ No CP |
| Radovic & Badawy, 2020^75^ | Review |
| Akalan et al., 2021^76^ | No NF intervention |

| ***cont.* Appendix S2: Excluded references in English and non-English language** | | | | | |
| --- | --- | --- | --- | --- | --- |
| ***Non-English language references excluded after full-text review (n=2) or abstract screening (n=13)*** | | | | | |
| Study | Original language | Abstract available in English  (Yes/No) | Full text translation  (N/A; Google Translate) | Exclusion level  (AS; FT) | Reason for exclusion |
| Pfister, 2018^77^ | German | Yes | Google Translate | FT | Review; deep brain stimulation |
| Jimenez et al., 2024^78^ | Spanish | Yes | Google Translate | FT | No NF intervention; design of a wheelchair |
| Gavrilovic, 2022^79^ | Serbian | Yes | N/A | AS | Thesis; human gait |
| Katic, 2023^80^ | Serbian | Yes | N/A | AS | Thesis; neuroprosthetic feedback |
| Guillen et al., 2023^81^ | Spanish | Yes | N/A | AS | Systematic review; self-regulated learning |
| Abusueva et al., 2023^82^ | Russian | Yes | N/A | AS | No NF intervention; comorbidity in autism |
| Mete, 2024^83^ | Turkish | Yes | N/A | AS | Thesis; stroke |
| Ismayıllı, 2023^84^ | Azerbaijani | Yes | N/A | AS | Thesis; depression |
| Carvoeiras, 2023^85^ | Portuguese | Yes | N/A | AS | Thesis; privacy software |
| Wang et al., 2019^86^ | Chinese | Yes | N/A | AS | No NF intervention/ No CP; emotions |
| Wang et al., 2005^87^ | Chinese | Yes | N/A | AS | No NF intervention/ No CP; sleep |
| Faber et al., 2001^88^ | Czech | Yes | N/A | AS | No CP; ADHD |
| Pastor & Balana, 2002^89^ | Spanish | Yes | N/A | AS | Review; biofeedback |
| Santos et al., 2018^90^ | Portuguese | Yes | N/A | AS | No NF intervention; virtual reality |
| Kubik, 2010^91^ | Polish | Yes | N/A | AS | Review; NF |
| N/A, Not Applicable – translation was not necessary for papers with available abstract in English language; AS, paper was excluded after abstract screening; FT, paper was excluded after full text review; NF, neurofeedback; CP, cerebral palsy; ADHD, attention deficit and hyperactivity disorder | | | | | |

***References only for Supporting material Appendix S2***

10.	Floreani ED, Rowley D, Kelly D, Kinney-Lang E, Kirton A. On the feasibility of simple brain-computer interface systems for enabling children with severe physical disabilities to explore independent movement. Frontiers in Human Neuroscience. 2022.

11.	Gad P, Hastings S, Zhong H, Seth G, Kandhari S, Edgerton VR. Transcutaneous Spinal Neuromodulation Reorganizes Neural Networks in Patients with Cerebral Palsy. Neurotherapeutics. 2021;18(3):1953-62.

12.	Girshin K, Sachdeva R, Cohn R, Gad P, Krassioukov AV, Edgerton VR. sPinal cOrd neUromodulatioN to treat Cerebral palsy in pEdiatrics: POUNCE Multisite Randomized Clinical Trial. Frontiers in Neuroscience. 2023;17.

38.	Syam SH-F, Lakany H, Ahmad RB, Conway BA. Comparing Common Average Referencing to Laplacian Referencing in Detecting Imagination and Intention of Movement for Brain Computer Interface. Les Ulis: EDP Sciences PP - Les Ulis; 2017 2017.

39.	Vasiljevic GAM, Cunha de Miranda L. The CoDIS Taxonomy for Brain-Computer Interface Games Controlled by Electroencephalography. International Journal of Human-Computer Interaction. 2023.

40.	Zahradka N, Behboodi A, Sansare A, Lee SCK. Evaluation of individualized functional electrical stimulation-induced acute changes during walking: A case series in children with cerebral palsy. Sensors. 2021;21(13).

41.	Ramakrishnan J. Cerebral palsy-affected individuals' brain-computer interface for wheelchair movement in an indoor environment using mental tasks. EGYPTIAN INFORMATICS JOURNAL. 2024;26.

42.	An Y, Min S, Park C. Clinical effects of a novel deep learning-based rehabilitation application on cardiopulmonary function, dynamic and static balance, gait function, and activities of daily living in adolescents with hemiplegic cerebral palsy. Medicine. 2024;103(10):1.

43.	Jadavji Z, Kirton A, Metzler MJ, Zewdie E. BCI-activated electrical stimulation in children with perinatal stroke and hemiparesis: A pilot study. Frontiers in Human Neuroscience. 2023;17.

51.	Chen W, Liu X, Wan P, Chen Z, Chen Y. Anti-artifacts techniques for neural recording front-ends in closed-loop brain-machine interface ICs. Frontiers in Neuroscience. 2024;18.

52.	Chung-Min W, Chen Y-J, Shih-Chung C, Sheng-Feng Z. Creating an AI-Enhanced Morse Code Translation System Based on Images for People with Severe Disabilities. Bioengineering. 2023;10(11):1281.

53.	Dion K, Rowley D, Floreani ED, Kinney-Lang E, Robu I, Kirton A. Think BIG: Brain-Computer Interface Goals for Children with Quadriplegic Cerebral Palsy. Piscataway: The Institute of Electrical and Electronics Engineers, Inc. (IEEE); 2023.

62.	Moufassih M, Tarahi O, Hamou S, Agounad S, Idrissi Azami H. Boosting motor imagery brain-computer interface classification using multiband and hybrid feature extraction. Multimedia Tools and Applications. 2023.

63.	Raj U, Mukul MK. Home Automation Using Brain–Computer Interface2023 2023. 639-49 p.

64.	Ranzani R, Razzoli M, Sanson P, Song J, Galati S, Ferrarese C, et al. Feasibility of Adjunct Therapy with a Robotic Hand Orthosis after Botulinum Toxin Injections in Persons with Spasticity: A Pilot Study. Toxins. 2024;16(8):346.

65.	Saltzman H, Rajaram R, Zhang Y, Islam MA, Kiourti A. Wearable Loops for Dynamic Monitoring of Joint Flexion: A Machine Learning Approach. Electronics. 2024;13(12):2245.

66.	Sawai S, Murata S, Fujikawa S, Yamamoto R, Shima K, Nakano H. Effects of neurofeedback training combined with transcranial direct current stimulation on motor imagery: A randomized controlled trial. Frontiers in Neuroscience. 2023.

67.	Siddiqui F, Awwab M, Alam MA, Naaz S, Agarwal P, Shahab Saquib S, et al. Deep Neural Network for EEG Signal-Based Subject-Independent Imaginary Mental Task Classification. Diagnostics. 2023;13(4):640.

68.	Sreeja SR, Samanta D. Dictionary reduction in sparse representation-based classification of motor imagery EEG signals. Multimedia Tools and Applications. 2023;82(20):31157-80.

69.	Tou SLJ, Warschausky SA, Karlsson P, Huggins JE. Individualized Electrode Subset Improves the Calibration Accuracy of an EEG P300-design Brain-Computer Interface for People with Severe Cerebral Palsy. 2023.

79.	Gavrilović MM. Objektivizacija Ljudskog Hoda Primenom Metode Glavnih Komponenti Dobijenih Sa Signala Dinamike Stopala [Ph.D.]. Serbia: University of Belgrade (Serbia); 2023.

80.	Катић НM. Decoding Neural Mechanisms Using in Silico and Animal Models for Restoring Somatosensory Feedback with Neuroprostheses [Ph.D.]. Serbia: University of Belgrade (Serbia); 2023.

81.	Guillén AL, Colón AMO, Moreno JR. Tecnologías inmersivas en el aprendizaje autorregulado: Revisión sistemática de literatura científica. Digital Education Review. 2023(44):105-13.

82.	Abusueva BA, Askevova MA, Shanavazova MD, Nasrutdinova BM, Bobylova MY, Gadzhieva AS. Comorbidity of cerebral palsy and epilepsy with autism in children. Russkii Zhunal Detskoi Nevrologii. 2023;18(2-3):14-21.

83.	Mete E. İnme Hastalarında Eşik Değerinde Elektrik Stimulasyonu ile Beraber Uygulanan Modifiye Kısıtlayıcı Zorunlu Hareket Tedavisinin Etkinliği [Ph.D.]. Turkey: Marmara Universitesi (Turkey); 2024.

84.	İsmayıllı A. Anadangəlmə Və Sonradan Qazanılmış Fiziki Məhdudiyyəti Olan Şəxslərdə Depressiyanın Müqayisəli Təhlili [Master's]. Azerbaijan: Khazar University (Azerbaijan); 2023.

85.	Carvoeiras FJdEPF. Proteção de ficheiros de log do software eugénio Desenvolvimento de mecanismos de segurança e privacidade [Master's]. Portugal: Instituto Politecnico de Beja (Portugal); 2023.
