## Supplementary material for "Effectiveness of neurofeedback interventions in cerebral palsy: A systematic review": The ROBINS-I V2 assessment is shown in Appendix S3

| **Appendix S3. Risk of Bias In Non-Randomized Studies – of Interventions (ROBINS-I V2)^1^** | | | | | | | | |
| --- | --- | --- | --- | --- | --- | --- | --- | --- |
| **Study** | **Pre-intervention** | | **At intervention** | **After intervention** | | | |  |
|  | **Confounding** | **Selection of participants** | **Classification of intervention** | **Deviation from intended interventions** | **Missing data** | **Measurement of outcomes** | **Selection of reported results** | **Overall risk of bias assessment** |
| Alves-Pinto et al., 2017^2^ | Serious | Low | Low | Low | Low | Serious | Serious | Critical |
| Larina et al., 2020^3^ | Serious | Low | Low | Moderate | Critical | Moderate | Low | Critical |
| Taherian et al., 2015^4^ | Serious | Low | Low | Moderate | Serious | Moderate | Low | Critical |
| Pavlenko et al., 2024^5^ | Serious | Low | Moderate | Moderate | Low | Moderate | Low | Serious |
| Xie et al.,2021^6^ | Serious | Low | Low | Low | Moderate | Low | Serious | Critical |
| Pavlenko et al., 2023^7^ | Critical | Serious | Low | Serious | Moderate | Moderate | Low | Critical |
| Pavlenko et al., 2024^8^ | Serious | Low | Low | Low | Low | Moderate | Low | Serious |
| **Moderate risk of bias** indicates that the study is ‘sound for a non-randomized study’ but not comparable to a rigorous randomized trial; **Serious risk of bias** indicates the presence of ‘important problems’; **Critical risk of bias** indicated the study is ‘too problematic to provide any useful evidence on the effects of intervention’ | | | | | | | | |

**References: (Appendix S3 only)**
