## Supplementary material for "Effectiveness of neurofeedback interventions in cerebral palsy: A systematic review": the RoB 2 in Appendix S4

| **Appendix S4. Revised Cochrane risk-of-bias tool for randomized trials (ROB 2)^1^** | | | | | | |
| --- | --- | --- | --- | --- | --- | --- |
| **Study** | **Pre-intervention** | **After intervention** | | | |  |
|  | **Randomization process** | **Deviation from intended interventions** | **Missing outcome data** | **Measurement of outcomes** | **Selection of reported results** | **Overall risk of bias assessment** |
| Gupta et al., 2020^2^ | Some concerns | Some concerns | Low | Some concerns | Some concerns | High |
| Kim & Lee, 2016^3^ | Low | Some concerns | Low | Some concerns | Some concerns | High |
| Korendah et al., 2021^4^ | Some concerns | Some concerns | Low | Some concerns | Some concerns | High |
| Yu et al., 2012^5^ | Some concerns | Some concerns | Low | Some concerns | Some concerns | High |
| Larina et al., 2019^6^ | Low | Some concerns | High | High | High | Some concerns |
| Korsunskaya et al., 2020^7^ | Some concerns | High | Low | Some concerns | Some concerns | High |
| Chen et al., 2024^8^ | Low | High | High | Low | Some concerns | Some concerns |

**References: (Appendix S4 only)**

1.	Sterne JAC, Savović J, Page MJ, Elbers RG, Blencowe NS, Boutron I, et al. RoB 2: a revised tool for assessing risk of bias in randomised trials. BMJ. 2019;366:l4898.

2.	Gupta M, Bhatia D, Sinha TK, Dogra S, Jha SK. Investigation of cognitive changes in cerebral palsy children employing different integrated sensing techniques. Sensors International. 2020;1.
