## Supplementary material for "Effectiveness of neurofeedback interventions in cerebral palsy: A systematic review": and the SCD risk of bias tool in Appendix S5.

| **Appendix S5. Single-case design risk of bias (SCD RoB) tool^1^** | | | | | | | | |
| --- | --- | --- | --- | --- | --- | --- | --- | --- |
|  | **Selection Bias** | | **Performance Bias** | | **Detection Bias** | | | |
| Study | Sequence generation | Participant Selection | Blinding of participants and personnel | Procedural fidelity | Blinding of outcome assessors | Selective outcome reporting | Dependent variable reliability | Data sampling |
| Filho et al., 2017^2^ | H | L | H | H | H | L | H | H |
| Legarda et al., 2011^3^ | H | L | H | H | H | L | H | H |
| Neuper et al., 2003^4^ | H | L | H | L | H | L | L | L |
| Taherian et al., 2016^5^ | H | L | H | L | H | L | L | L |
| L = low risk of bias  U = unclear risk of bias  H = high risk of bias | | | | | | | | |

| **Type of bias** | **Definition** | **Domains** | **Description** |
| --- | --- | --- | --- |
| Selection bias | The systematic differences between baseline characteristics of the participants compared. | Sequence generation | The procedures used to allocate participants to intervention conditions or the order of the conditions to which participants are exposed. |
|  |  | Participant selection | The criteria and process used to include and select participants appropriate for the research. |
| Performance bias | The systematic differences between participants in the care provided or in the exposure to factors other than the intervention under investigation. | Blinding of participants and personnel | The methods used to ensure members of the research team remain unaware of when the intervention is implemented to whom. |
|  |  | Procedural fidelity | The quality of the description for each experimental condition and the reporting of evidence indicating sufficient adherence to the intervention under investigation. This includes the procedures used to train implementers. |
| Detection bias | The systematic differences between participants in how outcomes are determined. | Blinding of outcome assessors | The procedures used to ensure the individuals collecting outcome data are unaware of the study conditions and research purpose. |
|  |  | Selective outcome reporting | The completeness of the data reported for all participants who began the study including those who withdrew and for each of the dependent variables. |
|  |  | Dependent variable reliability | The procedures and reporting of agreement or reliability indices for the outcome variables. |
|  |  | Data sampling | The extent to which the amount data collected for the research was sufficient to determine the level and trend of the data patterns in each condition to support the determination of a functional relation |

**References: (Appendix S5 only)**
