## Supplementary material for "Effectiveness of neurofeedback interventions in cerebral palsy: A systematic review": The Downs and Black checklist is reported in Appendix S6,

| **Appendix S6. Assessment of the methodological quality based on Downs and Black checklist^1^** | | | | | | | | | | | | | | | | | | | | | | | | | | | |
| --- | --- | --- | --- | --- | --- | --- | --- | --- | --- | --- | --- | --- | --- | --- | --- | --- | --- | --- | --- | --- | --- | --- | --- | --- | --- | --- | --- |
| **Study** | **Items** | | | | | | | | | | | | | | | | | | | | | | | | | | |
|  | **Reporting** | | | | | | | | | | **External Validity** | | | **Internal validity-bias** | | | | | | | **Internal validity – confounding (selection bias)** | | | | | | **Power** |
|  | **Q1** | **Q2** | **Q3** | **Q4** | **Q5** | **Q6** | **Q7** | **Q8** | **Q9** | **Q10** | **Q11** | **Q12** | **Q13** | **Q14** | **Q15** | **Q16** | **Q17** | **Q18** | **Q19** | **Q20** | **Q21** | **Q22** | **Q23** | **Q24** | **Q25** | **Q26** | **Q27** |
| Alves-Pinto et al.,2017 ^2^ | 1 | 1 | 1 | 1 | 1 | 1 | 1 | 0 | 0 | 0 | 1 | 0 | 0 | 1 | 0 | 1 | 1 | 1 | 1 | 1 | 1 | 0 | 0 | 0 | 0 | u.t.d. | 1 |
| Gupta et al., 2020^3^ | 1 | 1 | 1 | 0 | 0 | 1 | 1 | 0 | 0 | 1 | 1 | 0 | 0 | 0 | 0 | 1 | 1 | 1 | 1 | 1 | 1 | 0 | 1 | 0 | 0 | u.t.d. | 1 |
| Kim & Lee, 2016^4^ | 1 | 1 | 1 | 1 | 1 | 1 | 1 | 0 | 1 | 1 | 1 | 0 | 1 | 0 | 0 | 1 | 1 | 1 | 1 | 1 | 1 | 1 | 1 | 0 | 0 | u.t.d. | 1 |
| Larina et al., 2020^5^ | 1 | 1 | 1 | 1 | 1 | 1 | 1 | 0 | 0 | 0 | 1 | 0 | 1 | 0 | 0 | 1 | 1 | 1 | 1 | 1 | 1 | 1 | 0 | 0 | 0 | u.t.d. | 1 |
| Taherian et al., 2015^6^ | 1 | 1 | 1 | 1 | 0 | 1 | 0 | 0 | 1 | 0 | 0 | 0 | 1 | 0 | 0 | 1 | 0 | 0 | 0 | 0 | u.t.d. | u.t.d. | 0 | 0 | 0 | u.t.d. | 0 |
| Korendah et al., 2021^7^ | 1 | 1 | 1 | 1 | 1 | 1 | 0 | 0 | 0 | 1 | 0 | 0 | 1 | 0 | 0 | 1 | 1 | 1 | 1 | 1 | 1 | 1 | 1 | 0 | 0 | u.t.d. | 1 |
| Yu et al., 2012^8^ | 1 | 1 | 1 | 1 | 1 | 1 | 1 | 0 | 1 | 1 | 0 | 0 | 1 | 0 | 0 | 1 | 1 | 1 | 1 | 1 | 1 | 1 | 1 | 0 | 0 | u.t.d. | 1 |
| Larina et al., 2019^9^ | 1 | 1 | 1 | 1 | 0 | 1 | 1 | 0 | 0 | 0 | 1 | 0 | 1 | 0 | 0 | 1 | 1 | 1 | 1 | 1 | 1 | 1 | 1 | 0 | 0 | u.t.d. | 1 |
| Xie et al., 2021^10^ | 1 | 1 | 1 | 1 | 0 | 1 | 1 | 0 | 0 | 1 | 1 | 0 | 0 | 0 | 0 | 0 | 1 | 1 | 0 | 1 | 0 | 0 | 0 | 0 | 0 | u.t.d. | 1 |
| Korsunskaya et al., 2020^11^ | 1 | 1 | 1 | 1 | 1 | 1 | 1 | 0 | 0 | 1 | 0 | 0 | 1 | 0 | 0 | 1 | 1 | 1 | 1 | 1 | 1 | 1 | 1 | 0 | 0 | u.t.d. | 1 |
| Pavlenko et al., 2023^12^ | 1 | 1 | 1 | 1 | 0 | 1 | 1 | 0 | 0 | 1 | 0 | 0 | 1 | 0 | 0 | 1 | 1 | 1 | 1 | 1 | 1 | 1 | 0 | 0 | 0 | u.t.d. | 1 |
| Pavlenko et al., 2024^13^ | 1 | 1 | 1 | 1 | 0 | 1 | 1 | 0 | 0 | 1 | 0 | 0 | 1 | 0 | 0 | 1 | 1 | 1 | 1 | 1 | 1 | 1 | 0 | 0 | 0 | u.t.d. | 1 |
| Pavlenko et al., 2024^14^ | 1 | 1 | 1 | 1 | 0 | 1 | 1 | 0 | 0 | 1 | 0 | 0 | 0 | 0 | 0 | 0 | 1 | 1 | 1 | 1 | 1 | 0 | 0 | 0 | 0 | u.t.d. | 1 |
| Chen et al., 2024 ^15^ | 1 | 1 | 1 | 1 | 1 | 1 | 1 | 0 | 1 | 1 | 0 | 0 | 0 | 1 | 0 | 0 | 1 | 1 | 1 | 1 | 1 | 1 | 1 | 0 | 0 | u.t.d. | 1 |
| u.t.d., unable to determine | | | | | | | | | | | | | | | | | | | | | | | | | | | |

| *Continued* Appendix S6. Assessment of the methodological quality based on Downs and Black checklist | | |
| --- | --- | --- |
| **Study** | **Total score** | **Quality level** |
|  |  | **Excellent: 26-32**  **Good: 20-25**  **Fair:15-19**  **Poor: ≤14** |
| Alves-Pinto et al.,2017 | 16 | Fair |
| Gupta et al., 2020 | 15 | Fair |
| Kim et al., 2016 | 20 | Good |
| Larina et al., 2020 | 17 | Fair |
| Taherian et al., 2015 | 7 | Poor |
| Korendah et al., 2021 | 17 | Fair |
| Yu et al., 2012 | 19 | Fair |
| Larina et al., 2019 | 17 | Fair |
| Xie et al., 2021 | 12 | Poor |
| Korsunskaya et al., 2020 | 18 | Fair |
| Pavlenko et al., 2023 | 16 | Fair |
| Pavlenko et al., 2024 | 16 | Fair |
| Pavlenko et al., 2024 | 13 | Poor |
| Chen et al., | 18 | Fair |
| u.t.d., unable to determine | | |

| **Items** |  | **Checklist question description** | | **Scoring key** |
| --- | --- | --- | --- | --- |
| ***Reporting*** | Q1 | Is the hypothesis/aim/objective of the study clearly described? |  | 1, yes;  0, no |
|  | Q2 | Are the main outcomes to be measured clearly described in the Introduction or Methods section? | If the main outcomes are first mentioned in the Results section, the question should be answered no. | 1, yes;  0, no |
|  | Q3 | Are the characteristics of the patients included in the study clearly described? | In cohort studies and trials, inclusion and/or exclusion criteria should be given. In case-control studies, a case-definition and the source for controls should be given. | 1, yes;  0, no |
|  | Q4 | Are the interventions of interest clearly described? | Treatments and placebo (where relevant) that are to be compared should be clearly described. | 1, yes;  0, no |
|  | Q5 | Are the distributions of principal confounders in each group of subjects to be compared clearly described? | A list of principal confounders is provided. | 2, yes;  1, partially;  0, no |
|  | Q6 | Are the main findings of the study clearly described? | Simple outcome data (including denominators and numerators) should be reported for all major findings so that the reader can check the major analyses and conclusions. (This question does not cover statistical tests which are considered below). | 1, yes;  0, no |
|  | Q7 | Does the study provide estimates of the random variability in the data for the main outcomes? | In non normally distributed data the inter-quartile range of results should be reported. In normally distributed data the standard error, standard deviation or confidence intervals should be reported. If the distribution of the data is not described, it must be assumed that the estimates used were appropriate and the question should be answered yes. | 1, yes;  0, no |
|  | Q8 | Have all important adverse events that may be a consequence of the intervention been reported? | This should be answered yes if the study demonstrates that there was a comprehensive attempt to measure adverse events. (A list of possible adverse events is provided). | 1, yes;  0, no |
|  | Q9 | Have the characteristics of patients lost to follow-up been described? | This should be answered yes where there were no losses to follow-up or where losses to follow-up were so small that findings would be unaffected by their inclusion.This should be answered no where a study does not report the number of patients lost to follow-up. | 1, yes;  0, no |
|  | Q10 | Have actual probability values been reported (e.g.0.035 rather than <0.05) for the main outcomes except where the probability value is less than 0.001? |  | 1, yes;  0, no |
| ***External validity*** | Q11 | Were the subjects asked to participate in the study representative of the entire population from which they were recruited? | The study must identify the source population for patients and describe how the patients were selected. Patients would be representative if they comprised the entire source population, an unselected sample of consecutive patients, or a random sample. Random sampling is only feasible where a list of all members of the relevant population exists. Where a study does not report the proportion of the source population from which the patients are derived,the question should be answered as unable to determine. | 1, yes;  0, no;  0, unable to determine |
|  | Q12 | Were those subjects who were prepared to participate representative of the entire population from which they were recruited? | The proportion of those asked who agreed should be stated. Validation that the sample was representative would include demonstrating that the distribution of the main confounding factors was the same in the study sample and the source population. | 1, yes;  0, no;  0, unable to determine |
|  | Q13 | Were the staff, places, and facilities where the patients were treated, representative of the treatment the majority of patients receive? | For the question to be answered yes the study should demonstrate that the intervention was representative of that in use in the source population. The question should be answered no if, for example, the intervention was undertaken in a specialist centre unrepresentative of the hospitals most of the source population would attend. | 1, yes;  0, no;  0, unable to determine |
| ***Internal validity - bias*** | Q14 | Was an attempt made to blind study subjects to the intervention they have received ? | For studies where the patients would have no way of knowing which intervention they received, this should be answered yes. | 1, yes;  0, no;  0, unable to determine |
|  | Q15 | Was an attempt made to blind those measuring the main outcomes of the intervention? |  | 1, yes;  0, no;  0, unable to determine |
|  | Q16 | If any of the results of the study were based on “data dredging”,was this made clear? | Any analyses that had not been planned at the outset of the study should be clearly indicated. If no retrospective unplanned subgroup analyses were reported, then answer yes. | 1, yes;  0, no;  0, unable to determine |
|  | Q17 | In trials and cohort studies, do the analyse sadjust for different lengths of follow-up of patients, or in case-control studies, is the time period between the intervention and outcome the same for cases and controls? | Where follow-up was the same for all study patients the answer should yes. If different lengths of follow-up were adjusted for by,for example, survival analysis the answer should be yes. Studies where differences in follow-up are ignored should be answered no. | 1, yes;  0, no;  0, unable to determine |
|  | Q18 | Were the statistical tests used to assess the main outcomes appropriate? | The statistical techniques used must be appropriate to the data. For example non-parametric methods should be used for small sample sizes. Where little statistical analysis has been undertaken but where there is no evidence of bias, the question should be answered yes. If the distribution of the data (normal or not) is not described it must be assumed that the estimates used were appropriate and the question should be answered yes. | 1, yes;  0, no;  0, unable to determine |
|  | Q19 | Was compliance with the intervention/s reliable? | Where there was non compliance with the allocated treatment or where there was contamination of one group, the question should be answered no. For studies where the effect of any misclassification was likely to bias any association to the null, the question should be answered yes. | 1, yes;  0, no;  0, unable to determine |
|  | Q20 | Were the main outcome measures used accurate (valid and reliable)? | For studies where the outcome measures are clearly described, the question should be answered yes. For studies which refer to other work or that demonstrates the outcome measures are accurate, the ques-tion should be answered as yes. | 1, yes;  0, no;  0, unable to determine |
| ***Internal validity - counfounding*** | Q21 | Were the patients in different intervention groups (trials and cohort studies) or were the cases and controls (case-control studies) recruited from the same population? | For example, patients for all comparison groups should be selected from the same hospital.The question should be answered unable to determine for cohort and case-control studies where there is no information concerning the source of patients included in the study. | 1, yes;  0, no;  0, unable to determine |
|  | Q22 | Were study subjects in different intervention groups (trials and cohort studies) or were the cases and controls (case-control studies) recruited over the same period of time? | For a study which does not specify the time period over which patients were recruited, the question should be answered as unable to determine. | 1, yes;  0, no;  0, unable to determine |
|  | Q23 | Were study subjects randomised to intervention groups? | Studies which state that subjects were randomised should be answered yes except where method of randomisation would not ensure random allocation. For example alternate allocation would score no because it is predictable. | 1, yes;  0, no;  0, unable to determine |
|  | Q24 | Was the randomised intervention assignment concealed from both patients and health care staff until recruitment was complete and irrevocable? | All non-randomised studies should be answered no. If assignment was concealed from patients but not from staff, it should be answered no. | 1, yes;  0, no;  0, unable to determine |
|  | Q25 | Was there adequate adjustment for confounding in the analyses from which the main findings were drawn? | This question should be answered no for trials if: the main conclusions of the study were based on analyses of treatment rather than intention to treat; the distribution of known confounders in the different treatment groups was not described; or the distribution of known confounders differed between the treatment groups but was not taken into account in the analyses. In non-randomised studies if the effect of the main confounders was not investigated or confounding was demonstrated but no adjust-ment was made in the final analyses the question should be answered as no. | 1, yes;  0, no;  0, unable to determine |
|  | Q26 | Were losses of patients to follow-up taken into account? | If the numbers of patients lost to follow-up are not reported, the question should be answered as unable to determine. If the proportion lost to follow-up was too small to affect the main findings, the question should be answered yes. | 1, yes;  0, no;  0, unable to determine |
| ***Power*** | Q27 | Did the study have sufficient power to detect a clinically important effect where the probability value for a difference being due to chance is less than 5%? | Sample sizes have been calculated to detect a difference of x% and y%. | Size of smallest intervention group:  5, n_8_+  4, n_7_-n_8_  3, n_5_-n_6_  2, n_3_-n_4_  1, n_1_-n_2_  0, <n_1_ |

**References: (Appendix S6 only)**

1.	Downs SH, Black N. The feasibility of creating a checklist for the assessment of the methodological quality both of randomised and non-randomised studies of health care interventions. Journal of Epidemiology and Community Health. 1998;52(6):377.

2.	Alves-Pinto A, Turova V, Blumenstein T, Hantuschke C, Lampe R. Implicit Learning of a Finger Motor Sequence by Patients with Cerebral Palsy After Neurofeedback. Appl Psychophysiol Biofeedback. 2017;42(1):27-37.

3.	Gupta M, Bhatia D, Sinha TK, Dogra S, Jha SK. Investigation of cognitive changes in cerebral palsy children employing different integrated sensing techniques. Sensors International. 2020;1.
