## Supplementary material for "Effectiveness of neurofeedback interventions in cerebral palsy: A systematic review": and SCED Scale is in Appendix S7.

| **Appendix S7. Single-Case Experimental Design (SCED) Scale^1^** | | | | | | | | | | | | |
| --- | --- | --- | --- | --- | --- | --- | --- | --- | --- | --- | --- | --- |
| Study | **Q1***  Clinical history | **Q2**  Target behaviors | **Q3**  Design | **Q4**  Baseline | **Q5**  Sampling behavior during treatment | **Q6**  Raw data recorded | **Q7**  Inter-rater reliability | **Q8**  Independence of assessors | **Q9**  Statistical analysis | **Q10**  Replication | **Q11**  Generalization | **Total score** |
| Filho et al., 2017^2^ | 1 | 1 | 0 | 1 | 1 | 1 | 1 | 0 | 0 | 0 | 0 | 5 |
| Legarda et al., 2011^3^ | 1 | 0 | 0 | 1 | 1 | 1 | 0 | 0 | 0 | 0 | 1 | 4 |
| Neuper et al., 2003^4^ | 1 | 1 | 1 | 1 | 1 | 1 | 1 | 0 | 1 | 0 | 0 | 7 |
| Taherian et al., 2016^5^ | 1 | 1 | 1 | 1 | 1 | 1 | 1 | 0 | 0 | 1 | 1 | 8 |
| Scores 0-4 = weak quality of evidence  Scores 5-7 = moderate quality of evidence  Scores 8-10 = high quality of evidence  *Q1 does not contribute to the total score. | | | | | | | | | | | | |

| **Descriptions of items in the Single-Case Experimental Design (SCED) Scale** | | |
| --- | --- | --- |
| **Item** | **Aim and brief definition** | **Scoring key** |
| Q1. Clinical history* | The study provides critical information regarding demographic and injury characteristics of the research subject that allows the reader to determine the applicability of the treatment to another individual. | 1, yes;  0, no |
| Q2. Target behaviors | The paper identifies a precise, repeatable and operationally defined target behavior that can be used to measure treatment success. | 1, yes;  0, no |
| Q3. Design | The study design allows the for the examination of cause-and-effect relationships to demonstrate treatment efficacy. | 1, yes;  0, no |
| Q4. Baseline | To establish that sufficient sampling of behavior had occurred during the pre-treatment period to provide an adequate baseline measure. | 1, yes;  0, no |
| Q5. Sampling behavior during treatment | To establish that sufficient sampling of behavior during the treatment phase has occurred to differentiate a treatment response from fluctuations in behavior that may have occurred at baseline. | 1, yes;  0, no |
| Q6. Raw data record | To provide an accurate representation of the variability of the target behavior. | 1, yes;  0, no |
| Q7. Inter-rater reliability | To determine if the target behavior measure is reliable and collected in a consistent manner. | 1, yes;  0, no |
| Q8. Independence of assessors | To reduce assessment bias by employing a person who is otherwise uninvolved in the study, to provide an evaluation of the patients. | 1, yes;  0, no |
| Q9. Statistical analysis | To demonstrate the effectiveness of the treatment of interest by statistically comparing the results over the study phases. | 1, yes;  0, no |
| Q10. Replication | To demonstrate that the application and results of the therapy are not limited to a specific individual or situation (i.e., that the results are reproduced in other circumstances – replicated across subjects, therapists or settings). | 1, yes;  0, no |
| Q11. Generalization | To demonstrate the functional utility of the treatment in extending beyond the target behaviours or therapy environment into other areas of the individual’s life. | 1, yes;  0, no |
| *Q1 does not contribute to the total score, i.e., the maximum SCED Scale score is 10/10. | | |

**References: (Appendix S7 only)**
